# Trans-biobank Mendelian randomization analyses identify opposing pathways in plasma low-density lipoprotein-cholesterol lowering and gallstone disease

**DOI:** 10.1101/2023.09.27.23296205

**Authors:** Guoyi Yang, Amy M Mason, Dipender Gill, C Mary Schooling, Stephen Burgess

**Author notes:** **Address for correspondence: Stephen Burgess**, Postal address: MRC Biostatistics Unit, University of Cambridge, East Forvie Building, Forvie Site, Robinson Way, Cambridge Biomedical Campus, Cambridge, UK, Tweeter handle: @stevesphd, **Guoyi Yang**, Postal address: School of Public Health, Li Ka Shing Faculty of Medicine, The University of Hong Kong, Hong Kong, China, Tweeter handle: @yanggy_hku. **Tweet:** Trans-biobank Mendelian randomization analyses support that different plasma LDL-cholesterol lowering pathways have distinct and opposing effects on risk of gallstone disease. Statins may reduce risk of gallstone disease. #LDL, #Gallstones, #Mendelian randomization.

## Abstract

**Background:** Plasma low-density lipoprotein (LDL)-cholesterol is positively associated with coronary artery disease risk while biliary cholesterol promotes gallstone formation.

**Objectives:** We tested the hypothesis that different plasma LDL-cholesterol lowering pathways have distinct effects on biliary cholesterol and thereby risk of gallstone disease.

**Methods:** This Mendelian randomization (MR) study used data from the UK Biobank (30,547 gallstone disease cases/336,742 controls), FinnGen (34,461 cases/301,383 controls) and Biobank Japan (9,305 cases/168,253 controls). First, drug-target MR and colocalization analyses were performed to investigate plasma LDL-cholesterol lowering therapies on gallstone disease. Second, clustered MR and pathway analyses were performed to identify distinct mechanisms underlying the association of plasma LDL-cholesterol with gallstone disease.

**Results:** For a 1-standard deviation reduction in plasma LDL-cholesterol, genetic mimics of statins were associated with lower risk of gallstone disease (odds ratio 0.72 [95% confidence interval 0.62, 0.83]) but PCSK9 inhibitors and mipomersen were associated with higher risk (1.11 [1.03, 1.19] and 1.23 [1.13, 1.35]). The association for statins was supported by colocalization (posterior probability 98.7%). Clustered MR analyses identified variant clusters showing opposing associations of plasma LDL-cholesterol with gallstone disease, with evidence for ancestry-and sex-specific associations. Among variants predicting lower plasma LDL-cholesterol, those associated with lower risk of gallstone disease were mapped to glycosphingolipid biosynthesis pathway, while those associated with higher risk were mapped to pathways relating to plasma lipoprotein assembly, remodelling, and clearance and ATP-binding cassette transporters.

**Conclusions:** Different plasma LDL-cholesterol lowering pathways may have opposing effects on risk of gallstone disease. Notably, statins may reduce risk of gallstone disease.

**Condensed abstract:** We hypothesized that different plasma LDL-cholesterol lowering pathways have distinct effects on risk of gallstone disease. We performed drug-target and clustered Mendelian randomization (MR) analyses, using data from the UK Biobank, FinnGen and Biobank Japan. Genetic mimics of statins were associated with lower risk of gallstone disease, but PCSK9 inhibitors and mipomersen were associated with higher risk. Clustered MR identified variant clusters showing opposing associations of plasma LDL-cholesterol with gallstone disease. This genetic study supports that different plasma LDL-cholesterol lowering pathways have opposing effects on risk of gallstone disease and statins may reduce risk of gallstone disease.

## Introduction

Cholesterol plays an important role in the aetiology of coronary artery disease (CAD) and gallstone disease. Plasma low-density lipoprotein (LDL)-cholesterol is positively associated with CAD risk (1), whereas biliary cholesterol promotes cholesterol gallstones formation (2). A recent randomized controlled trial (RCT) showed lowering plasma LDL-cholesterol with bempedoic acid, an adenosine triphosphate (ATP) citrate lyase inhibitor, increased risk of gallstones (3), but trial evidence for other lipid modifiers is limited (4). Therefore, it remains unclear whether the lithogenic effect is a general consequence of lowering plasma LDL-cholesterol or is unique to bempedoic acid.

RCTs are not usually designed or powered to identify adverse effects or novel indications. Observational studies suggest statin use is associated with lower gallstone disease risk (5,6), but these studies could be biased due to residual confounding or selection bias. Mendelian randomization (MR), an instrumental variable analysis with genetic instruments, is more robust to confounding than conventional observational studies (7). However, previous MR studies have yielded contradictory results, suggesting a positive (8) or null (9) association of lower plasma LDL-cholesterol with gallstone disease risk. Understanding the causal association of plasma LDL-cholesterol with gallstone disease has implications for repurposing plasma LDL-cholesterol lowering therapies and identifying potentially adverse effects.

Plasma LDL-cholesterol is regulated by different biological pathways (10), which could have distinct effects on biliary cholesterol and thereby gallstone disease. Statins reduce plasma LDL-cholesterol by inhibiting cholesterol biosynthesis (11), which may decrease biliary cholesterol (12) and reduce gallstone disease risk. However, pathways reducing plasma LDL-cholesterol while elevating biliary cholesterol, such as activating adenosine triphosphate (ATP)-binding cassette transporters G5/8 (ABCG5/8), may increase gallstone disease risk (13).

We hypothesized that different plasma LDL-cholesterol lowering pathways have distinct effects on risk of gallstone disease. First, we used MR to assess the associations of genetic mimics of current and emerging plasma LDL-cholesterol lowering therapies with gallstone disease risk. Second, we investigated distinct pathways underlying the association of plasma LDL-cholesterol with gallstone disease risk. Where possible, we assessed ancestry- and sex-specific associations, because gallstone prevalence is higher in European than Asian ancestry individuals, and in women than men (2).

## Methods

### Ethical approval

This study has been conducted using the UK Biobank Resource (Application number 98032). The UK Biobank obtained ethical approval from the North West Multi-centre Research Ethics Committee, and the participants provided written informed consent. The analysis of publicly available summary statistics does not require ethical approval.

### Study design

We used individual-level data from UK Biobank (14) and summary-level data from FinnGen (15) and Biobank Japan (16) for gallstone disease. We selected genetic mimics of plasma LDL-cholesterol lowering therapies from genes encoding the molecular target of each therapy. We conducted drug-target MR analyses to assess the associations of genetic mimics of each therapy with gallstone disease risk. We performed colocalization analyses to examine whether any associations found were driven by a shared causal variant between exposure and outcome or were confounded by linkage disequilibrium (17).

We extracted genetic predictors for plasma LDL-cholesterol from across the genome. We conducted clustered MR analyses to identify distinct clusters of genetic variants having similar causal estimates for plasma LDL-cholesterol on gallstone disease risk (18). We performed pathway analyses to investigate biological pathways relating to each cluster. A summary of the study design is shown in Supplemental Figure 1.

### Data sources

The UK Biobank recruited approximately 500,000 people (intended age 40-69 years, 94% self-reported European ancestry) between 2006 and 2010 from across the United Kingdom (14). Individual-level data used were under application 98032 (October 2021 updated). Cases were defined based on self-reported history of gallstones, International Classification of Diseases (ICD)-9 and ICD-10 codes related to gallstones, and medical treatment of gallstones, as previously (19). Both prevalent and incident cases were included. Controls were individuals without gallstone-related disease or treatment. Individuals who underwent cholecystectomy due to an alternative pathology (e.g., neoplasm) were excluded. We included 367,289 unrelated individuals of European ancestry (cases = 21,201 women/9,346 men, controls = 177,478 women/159,264 men) with genomic data passing quality control as described previously (20). We used logistic regression to obtain sex-combined and sex-specific genetic associations with gallstone disease.

Summary-level data from FinnGen (R8 release) included 34,461 cases and 301,383 controls (mean age 52 years, 55.7% women) (15). Cases were defined based on ICD-8, 9, 10 codes related to gallstones (15). Summary-level data from Biobank Japan included 9,305 cases and 168,253 controls (mean age 63 years, 46.3% women) (16). Cases were defined based on ICD-10 codes (16). Detailed definitions of gallstone disease are provided in Supplemental Table 1.

### Genetic instruments

We used ancestry-specific summary-level data from the Global Lipids Genetics Consortium (GLGC) for plasma LDL-cholesterol (1,231,289/82,587 people of European/East Asian ancestry) (21). We selected genetic mimics of plasma LDL-cholesterol lowering therapies from genes encoding the molecular targets of each therapy (i.e., *HMGCR* for statins, *PCSK9* for proprotein convertase subtilisin/kexin type 9 (PCSK9) inhibitors, *NPC1L1* for ezetimibe, *ACLY* for ATP citrate lyase inhibitors, *LDLR* for targeting LDL receptors, and *APOB* for mipomersen). We used genetic mimics of targeting ABCG5/8 from *ABCG5/8* as a positive control exposure, because *ABCG5/8* is a well-established lithogenic gene (2). We included all variants within 100kb on either side of each target gene that were in low linkage disequilibrium (r^2^ <0.1) and were genome-wide significantly (*p* value <5×10^-8^) associated with plasma LDL-cholesterol. We used a less stringent cut-off for linkage disequilibrium (r^2^ <0.1) to obtain more variants in each gene region to increase the power (22). We also extracted genetic predictors for plasma LDL-cholesterol from across the genome that were uncorrelated (r^2^ <0.001) and genome-wide significantly (*p* value <5×10^-8^) associated with plasma LDL-cholesterol. Estimates were expressed in 1-standard deviation (around 0.87 mmol/L) reduction in plasma LDL-cholesterol.

We used the F-statistic to assess instrument strength, approximated by the square of each SNP-exposure association divided by the square of its standard error (23). We used PhenoScanner, a database of genotype-phenotype associations (24,25), to check whether SNPs were genome-wide significantly (*p* value <5×10^-8^) associated with common confounders (i.e., socioeconomic status, smoking, alcohol drinking and physical activity). We included these SNPs in the main analysis, and excluded them in the sensitivity analysis. We also used positive control outcomes, i.e., CAD from CARDIoGRAMplusC4D Consortium (60,801 cases and 123,504 controls) (26) for people of European ancestry and myocardial infarction (MI) from Biobank Japan (14,992 cases and 146,214 controls) (16) for East Asians. A summary of genome-wide association studies (GWAS) used is provided in Supplemental Table 2.

### MR analysis

We aligned SNPs based on alleles and allele frequencies. We used proxy SNPs (r^2^≥0.8), where possible, when SNPs were not available in the outcome GWAS. We calculated MR estimates by meta-analyzing Wald estimates (the ratio of the genetic association with outcome to the genetic association with exposure) using inverse variance weighting (IVW) with fixed effects for three SNPs or fewer and random effects for four SNPs or more (27). To assess the robustness of the IVW estimates, we conducted sensitivity analyses using weighted median (28) and MR Egger (29). For the SNPs in low linkage disequilibrium (r^2^ <0.1), we calculated IVW and MR Egger estimates taking into account their correlations.

We meta-analyzed MR estimates from the three biobanks using a fixed-effects model unless the Q-statistic suggested heterogeneity when random effects were used. We assessed differences by sex using a two-sided z-test (30).

### Colocalization analysis

We performed colocalization analyses in a Bayesian framework to assess the posterior probability of a shared variant associated with both plasma LDL-cholesterol and gallstone disease (17). We included variants (minor allele frequency >0.1%) in or near (+/-100kb) the target gene where any associations were identified. A posterior probability larger than 0.80 provides evidence for colocalization (17). We set the prior probabilities as recommended, i.e., 1.0e-4 for a variant associated with plasma LDL-cholesterol, 1.0e-4 for a variant associated with gallstone disease, and 1.0e-5 for a variant associated with both traits (17). We also calculated the posterior probability for a shared variant associated with both traits conditional on the presence of a variant associated with gallstone disease, as the power to detect colocalization is low when the variants are not strongly associated with the outcome (31).

### Clustered MR

We used the MR-Clust method to identify distinct clusters of genetic variants having similar causal estimates for plasma LDL-cholesterol on gallstone disease, which might reflect distinct biological pathways (18). The MR-Clust accounts for differential uncertainty in the causal estimates, and includes a null cluster where SNP-specific estimates are centred around zero and a junk cluster where SNP-specific estimates are highly dispersed and are considered outliers (18). The presence of null and junk clusters requires substantial evidence of similarity to define a cluster, which avoids the detection of spurious clusters (18). We only included variants with inclusion probability >0.80 in each cluster, and only reported a cluster if at least four variants satisfy this criterion, as recommended (18).

### Pathway analysis

We performed pathway analysis to examine biological pathways relating to each variant cluster using the Functional Mapping and Annotation (FUMA) platform (32). We first applied the SNP2GENE function to map cluster-specific genetic variants to genes, where we used 100-kb positional mapping and expression quantitative trait locus (eQTL) mapping based on GTEx v8 (32). We then used the GENE2FUNC function to associate the mapped genes with biological pathways defined by KEGG and Reactome database (32).

All statistical analyses were conducted using R version 4.2.1 and the packages “ieugwasr”, “TwoSampleMR”, “MendelianRandomization”, “metafor”, “coloc”, and “mrclust”.

## Results

### Baseline characteristics of UK Biobank participants

Baseline characteristics of 367,289 UK Biobank participants (30,547 cases and 336,742 controls) included in this study are shown in Table 1. Cases were older at baseline (mean age 59.4 vs 57.0 years) and had higher body mass index (mean 29.6 vs 27.2 kg/m^2^) than controls. Cases had a greater proportion of women (69.4% vs 52.7%) and current lipid-lowering medication users (22.1% vs 16.5%) but had a smaller proportion of current alcohol drinkers (89.1% vs 93.6%) than controls.

**Table 1.** Baseline characteristics of gallstone disease cases and controls in the UK Biobank.

| Characteristics | Cases (N=30,547) | Controls (N=336,742) |
| --- | --- | --- |
| Age at recruitment, years | 59.4 (7.4) | 57.0 (8.0) |
| Sex |  |  |
| Men | 9,346 (30.6) | 159,264 (47.3) |
| Women | 21,201 (69.4) | 177,478 (52.7) |
| LDL-cholesterol, mmol/L | 3.5 (0.9) | 3.6 (0.9) |
| BMI, kg/m<sup>2</sup> | 29.6 (5.5) | 27.2 (4.6) |
| SBP, mmHg | 138.8 (18.4) | 137.6 (18.6) |
| Smoking |  |  |
| Current | 3,118 (10.2) | 34,712 (10.3) |
| Other | 27,429 (89.8) | 302,030 (89.7) |
| Alcohol drinking |  |  |
| Current | 27,204 (89.1) | 315,226 (93.6) |
| Other | 3,343 (10.9) | 21,516 (6.4) |
| Lipid-lowering medication |  |  |
| Current | 6,751 (22.1) | 55,467 (16.5) |
| Other | 23,796 (77.9) | 281,275 (83.5) |

BMI, body mass index; LDL, low-density lipoprotein; SBP, systolic blood pressure. Data are mean (standard deviation) for continuous variables or N (%) for categorical variables.

### Genetic instruments

We extracted SNPs for plasma LDL-cholesterol lowering therapies in people of European and East Asian ancestry, respectively (Supplemental Table 3). We did not extract SNPs for ATP citrate lyase inhibitors in people of European or East Asian ancestry, or for ezetimibe and targeting ABCG5/8 in East Asians, because the SNPs in or near the target genes were not genome-wide significantly associated with plasma LDL-cholesterol (*p* values >5×10^-8^). We extracted 324 and 43 SNPs for plasma LDL-cholesterol in people of European and East Asian ancestry, respectively (Supplemental Table 4).

The F-statistics for all SNPs were >10. Variance in plasma LDL-cholesterol explained by SNPs for each therapy and plasma LDL-cholesterol is provided in Supplemental Table 5. None of the SNPs for each therapy were genome-wide significantly associated with common confounders (*p* values >5×10^-8^), but four SNPs for plasma LDL-cholesterol were associated with smoking or alcohol drinking (Supplemental Table 6). As expected, SNPs for each therapy and lower plasma LDL-cholesterol were associated with lower risk of CAD and MI (Supplemental Figures 2-3).

### Drug-target MR

After meta-analyzing MR estimates from the three biobanks, genetic mimics of statins were associated with lower gallstone disease risk, while genetic mimics of PCSK9 inhibitors, mipomersen, and targeting ABCG5/8 were associated with higher gallstone disease risk (Figure 1). We did not observe heterogeneity in IVW estimates across biobanks (*p* values for heterogeneity >0.05). The weighted median and MR Egger gave similar interpretations (Figure 1). For a 1-standard deviation reduction in plasma LDL-cholesterol, associations for statins (odds ratio for women 0.76 [95% confidence interval 0.58, 1.00] vs men 0.99 [0.62 to 1.58]), PCSK9 inhibitors (women 1.09 [0.95 to 1.26] vs men 1.36 [1.11 to 1.67]) and targeting ABCG5/8 (women 243.40 [114.10 to 519.26] vs men 58.39 [29.94 to 113.86]) slightly differed by sex (Supplemental Figure 4, *p* values for sex differences 0.332, 0.081 and 0.006).

**Figure 1.**
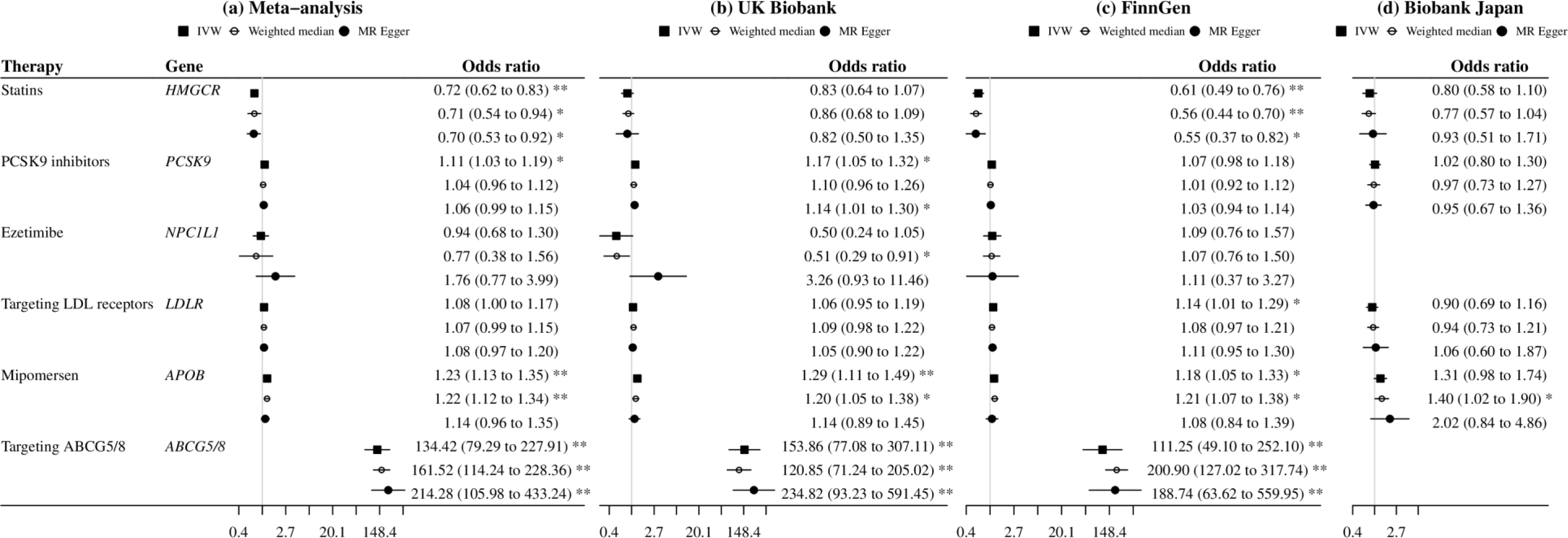
Mendelian randomization estimates for genetic mimics of plasma LDL-cholesterol lowering therapies on risk of gallstone disease. Estimates are expressed in odds ratio per 1-standard deviation (around 0.87 mmol/L) reduction in plasma LDL-cholesterol. * denotes *p* value <0.05; ** denotes *p* value <0.001.

### Colocalization analysis

Colocalization analyses were performed for plasma LDL-cholesterol with gallstone disease in or near (+-100kb) *HMGCR, PCSK9*, *APOB*, and *ABCG5/8* using the UK Biobank and FinnGen. The posterior probabilities for a shared variant associated with both traits were 98.7% for statins (rs12916 with the largest probability for both traits) but were <10% for the others (Figure 2). The posterior probabilities were >50% for a variant associated with plasma LDL-cholesterol only for PCSK9 inhibitors and mipomersen, and were >99.9% for independent variants associated with each trait for targeting ABCG5/8 (Supplemental Table 7).

**Figure 2.**
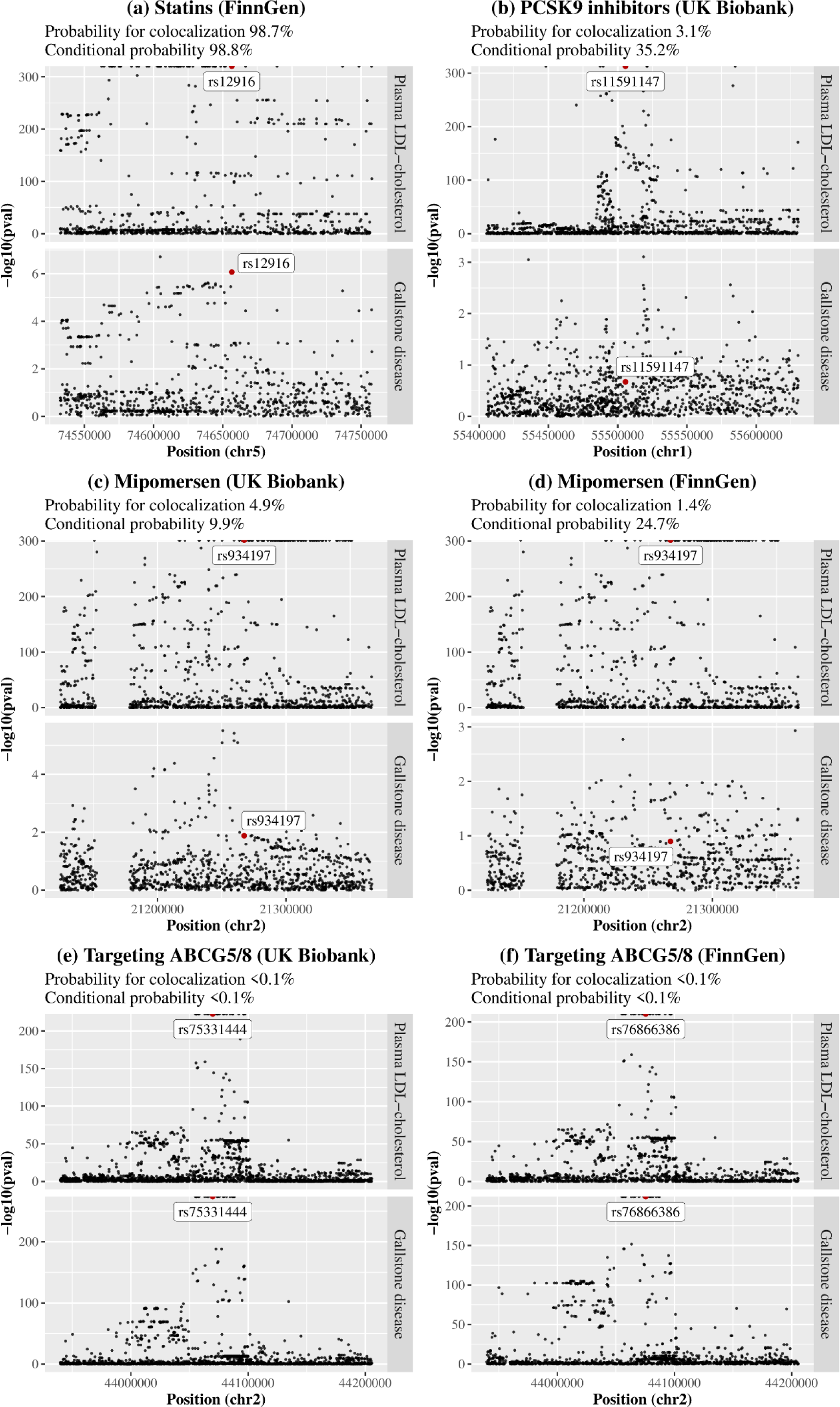
Colocalization analyses for plasma LDL-cholesterol with gallstone disease in or near (+-100kb) the target gene of each therapy. Prior probabilities were set to 1.0e-4 for a variant associated with plasma LDL-cholesterol, 1.0e-4 for a variant associated with gallstone disease, and 1.0e-5 for a variant associated with both traits. Probability for colocalization means the posterior probability for a shared variant associated with both traits; conditional probability means the posterior probability for a shared variant associated with both traits conditional on the presence of a variant associated with gallstone disease. The variant with the largest posterior probability for both traits is highlighted with a label.

### Clustered MR

Using all SNPs for lower plasma LDL-cholesterol, there was a positive association with gallstone disease risk in the UK Biobank and FinnGen but a null association in Biobank Japan (Figure 3). However, we observed heterogeneity in SNP-specific estimates (*p* values for heterogeneity <0.001). Clustered MR analyses consistently identified variant clusters showing opposing associations of lower plasma LDL-cholesterol with gallstone disease risk (Figures 3-4). By contrast, these clusters were generally associated with lower risk of CAD and MI (Supplemental Figure 3). Sensitivity analysis excluding SNPs associated with smoking or alcohol drinking did not change the results substantially (Supplemental Figures 5-6).

**Figure 3.**
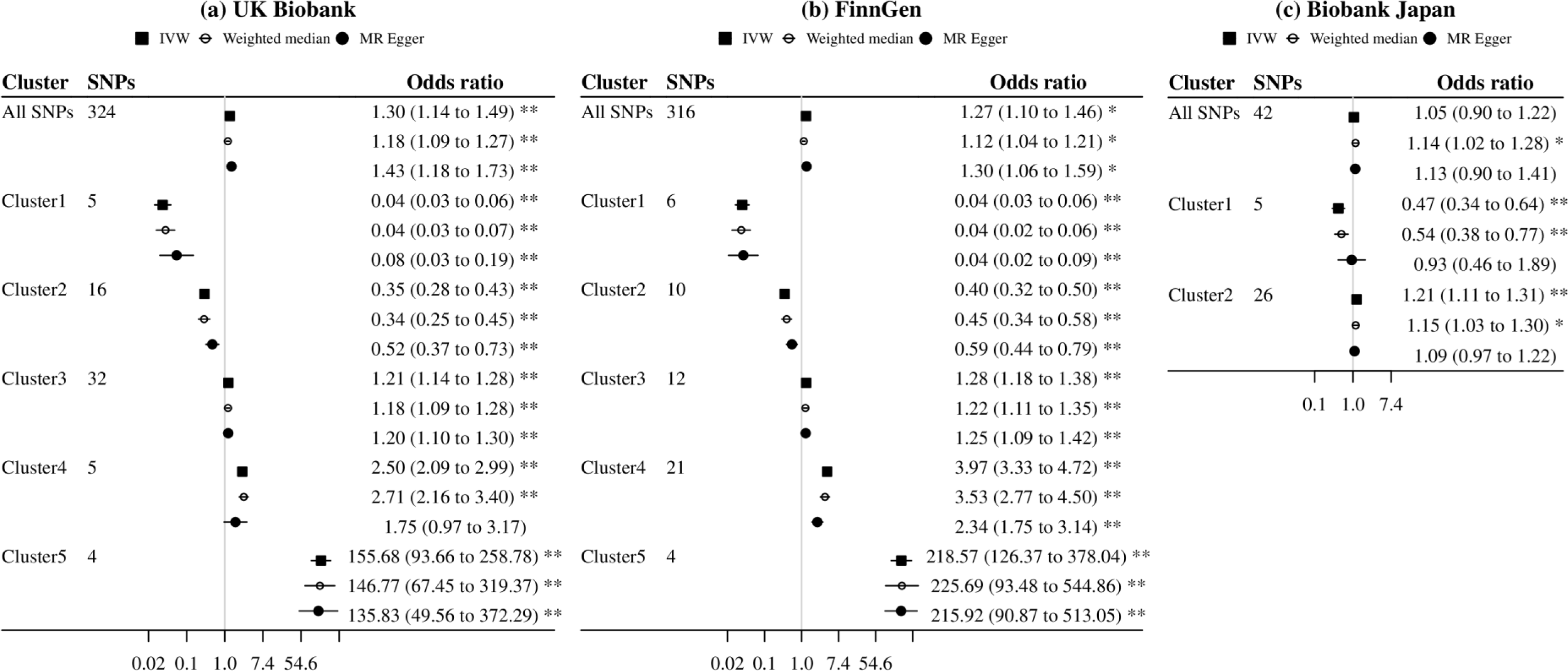
Mendelian randomization estimates for genetically predicted lower plasma LDL-cholesterol on risk of gallstone disease using all SNPs and cluster-specific SNPs (inclusion probability >0.80). Estimates are expressed in odds ratio per 1-standard deviation (around 0.87 mmol/L) reduction in plasma LDL-cholesterol. * denotes *p* value <0.05; ** denotes *p* value <0.001.

**Figure 4.**
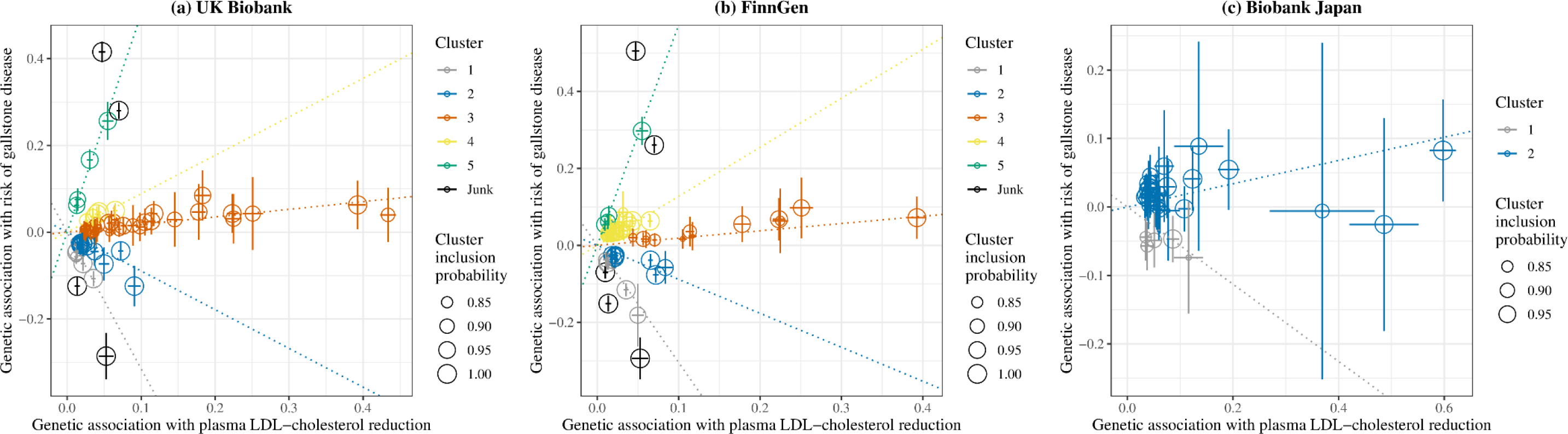
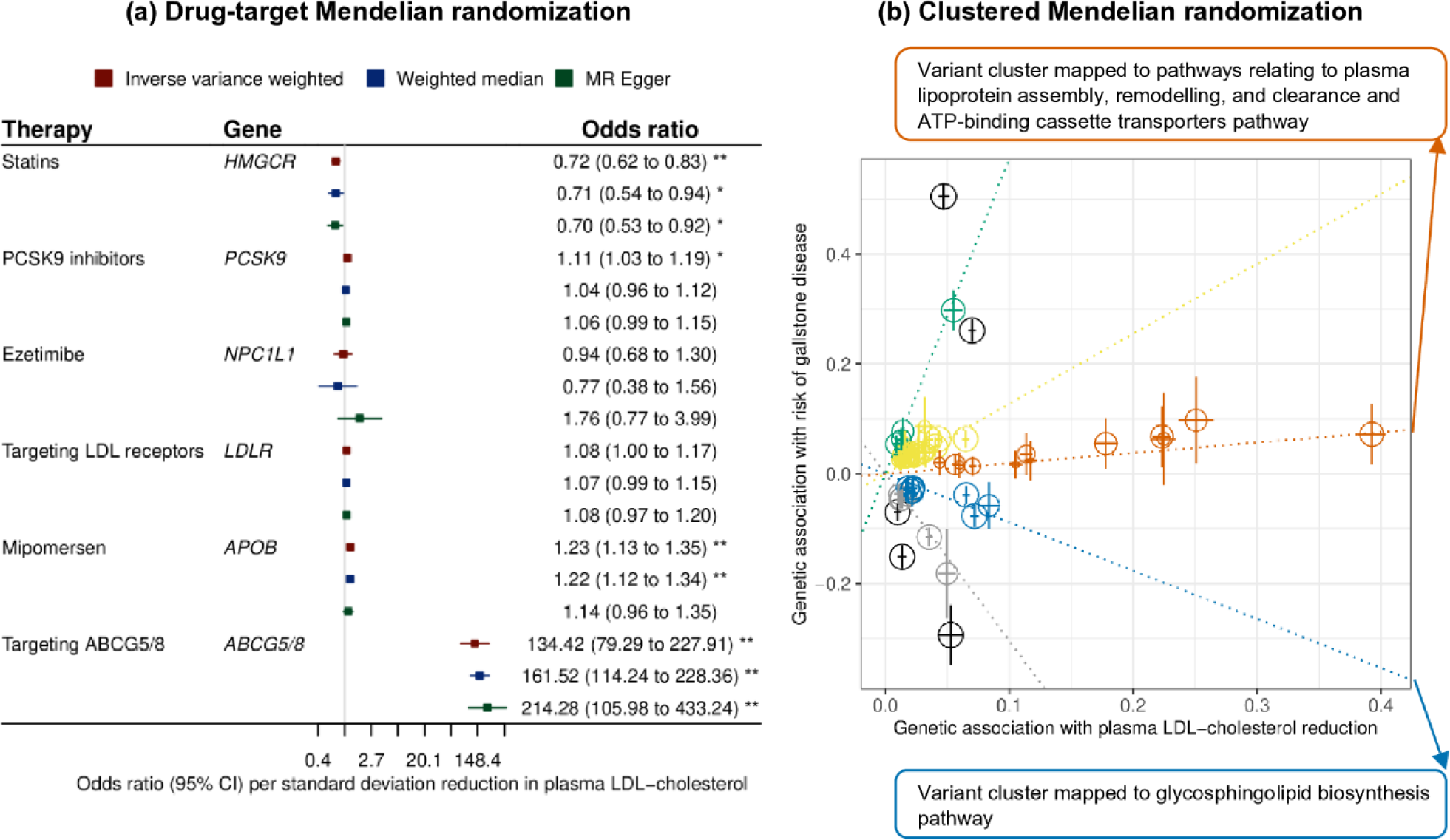
Genetic associations with plasma LDL-cholesterol reduction (standard deviation) and risk of gallstone disease (log odds) for SNPs with inclusion probability >0.80 in clustered Mendelian randomization analyses. Points represent SNPs; dotted lines are cluster means; error bars are 95% confidence intervals for genetic associations. Central Illustration. Drug-target and clustered Mendelian randomization analyses identify distinct and opposing pathways in the association of plasma LDL-cholesterol with gallstone disease. ABCG5/8, adenosine triphosphate (ATP)-binding cassette transporters G5/8; LDL, low-density lipoprotein; PCSK9, proprotein convertase subtilisin/kexin type 9. * denotes *p* value <0.05; ** denotes *p* value <0.001.

Cluster patterns were consistent in the UK Biobank and FinnGen (Figures 3-4), with overlapping cluster-specific SNPs (Supplemental Table 8) and similar SNP-specific estimates (Supplemental Figure 7). Sex-specific clustered MR analyses also showed such opposing associations, but the pattern appeared more evident in women than men (Supplemental Figures 8-9).

### Pathway analysis

We found clusters 2 and 3 in FinnGen, cluster 3 in the UK Biobank, and cluster 2 in Biobank Japan mapped to specific pathways (Supplemental Table 9). Cluster 2 in FinnGen showing an inverse association of lower plasma LDL-cholesterol with gallstone disease was mapped to glycosphingolipid biosynthesis pathway, while other clusters showing a positive association were mapped to pathways relating to plasma lipoprotein assembly, remodelling, and clearance and ATP-binding cassette transporters. Correspondingly, cluster 3 in FinnGen and UK Biobank and cluster 2 in Biobank Japan had similar estimates as mipomersen; cluster 5 in FinnGen and UK Biobank had similar estimates as targeting ABCG5/8 (Figures 1 and 3).

## Discussion

Consistent with previous observational studies (5,6), this study provides genetic evidence suggesting statins may reduce gallstone disease risk. Our investigation has added to the evidence base by identifying distinct and opposing pathways underlying the association of plasma LDL-cholesterol with gallstone disease.

Genetic evidence suggested statins may reduce the risk of gallstone disease, while PCSK9 inhibitors, mipomersen and targeting ABCG5/8 may increase the risk. These findings are consistent with observational studies showing long-term use of statins is associated with lower gallstone disease risk (5,6), and a previous MR study showing *ABCG5/8* variants lowering plasma LDL-cholesterol are associated with higher gallstone disease risk (13). However, an RCT showed simvastatin plus ezetimibe did not affect gallstone risk during a follow-up of 4.9 years (4), although the small number of gallstone cases may have limited the detection of any possible effect. Meta-analyses of RCTs showed statins reduced the risk of pancreatitis, a common complication of gallstone disease (33).

Plasma LDL-cholesterol lowering therapies may have distinct effects on biliary cholesterol and thereby gallstone disease. Statins decrease hepatic cholesterol synthesis (11), and may decrease biliary cholesterol (12) and facilitate cholesterol gallstone dissolution (34,35). PCSK9 inhibitors increase LDL receptors and mipomersen decreases apolipoprotein B-containing particles (11), which may increase hepatic and biliary cholesterol (2). Targeting ABCG5/8 inhibits cholesterol absorption and facilitates biliary cholesterol secretion (2). Increased biliary cholesterol accelerates supersaturation of bile and promotes cholesterol gallstone formation (2).

Colocalization analysis substantiated the association of statins with gallstone disease. A lack of colocalization for PCSK9 inhibitors and mipomersen is possibly due to insufficient power. However, colocalization analysis suggested the association of targeting ABCG5/8 with gallstone disease was confounded by linkage disequilibrium. This could be explained by different lead variants for plasma versus biliary cholesterol in or near *ABCG5/8*. Such differences would also explain the implausibly high MR estimates for targeting ABCG5/8, which are presented in effect sizes of plasma LDL-cholesterol reduction.

Using all SNPs for lower plasma LDL-cholesterol, there was a positive or null association with gallstone disease, as in previous MR studies (8,9). However, we identified variant clusters showing opposing associations of plasma LDL-cholesterol with gallstone disease. Among variants predicting lower plasma LDL-cholesterol, those associated with lower gallstone disease risk were mapped to glycosphingolipid biosynthesis pathway, while those associated with higher gallstone disease risk were mapped to pathways relating to plasma lipoprotein assembly, remodelling, and clearance and ATP-binding cassette transporters. These findings are consistent with the evidence available and the mechanisms of plasma LDL-cholesterol lowering therapies. In vitro studies have showed statins affect glycosphingolipid profiles through inhibiting Rab prenylation (36), which could suppress gallstone formation (37). PCSK9 inhibitors and mipomersen are involved in plasma LDL assembly and clearance, and ABCG5/8 are key members of ATP-binding cassette transporters (11).

Unlike the effects on CAD or MI, the lithogenic effect is specific to certain pathways rather than a general consequence of lowering plasma LDL-cholesterol. Similarly, previous MR studies showed plasma LDL-cholesterol lowering therapies differed in their associations with body mass index (38) and type 2 diabetes (39). These insights have implications for identifying repurposing opportunities and adverse effects of plasma LDL-cholesterol lowering therapies.

The opposing associations of plasma LDL-cholesterol with gallstone disease seemed more evident in European than East Asian ancestry individuals and in women than men, consistent with different gallstone prevalence rates by ancestry and sex (2). Plasma LDL-cholesterol lowering therapies may have distinct effects on biliary cholesterol and thereby cholesterol gallstones; however, pigment gallstones are more common in East Asians than Europeans (40), which may explain the difference by ancestry. Statins partially operate through sex hormones (41), which could be relevant to the more marked association in women than men. Alternatively, statins decreasing calcium, another component of gallstones, specifically in women (42) might play a role.

### Study limitations

This trans-ethnic study takes advantage of three large-scale biobanks. The association of genetic mimics of statins with gallstone disease is novel, as are the distinct and opposing associations of plasma LDL-cholesterol with gallstone disease. This study has several limitations. First, MR relies on three rigorous assumptions, that is genetic instruments should be strongly related to the exposure, share no common cause with the outcome, and be independent of the outcome given the exposure (7). We calculated the F-statistics, checked genetic associations with common confounders, and used CAD and MI as positive control outcomes to assess the validity of genetic instruments. Second, we were unable to assess the lithogenic effect of ATP citrate lyase inhibitors due to a lack of valid instruments. Third, we performed colocalization analyses for plasma LDL-cholesterol with gallstone disease, while biliary cholesterol likely underlies any effects on gallstone disease. The discrepancy between plasma versus biliary cholesterol may partly explain some lack of colocalization. Replication using biliary cholesterol would be ideal when relevant GWAS becomes available. Fourth, not all the variant clusters could be mapped to specific pathways, possibly due to the small number of SNPs with inclusion probability >0.80 in some clusters. However, such a conservative inclusion criterion avoids the generation of spurious clusters by chance (18). Fifth, gallstone disease studied here is not specific to cholesterol gallstones and the definitions slightly vary across the three biobanks; however, we still observed consistent patterns. Finally, MR assesses lifelong effects, which cannot directly inform the quantitative effects of plasma LDL-cholesterol lowering therapies in the short term.

## Conclusions

This genetic study supports that different plasma LDL-cholesterol lowering pathways have distinct and opposing effects on risk of gallstone disease. Notably, statins may reduce risk of gallstone disease.

## Acknowledgements

The authors acknowledge the UK Biobank for approving our application, and GLGC, FinnGen, Biobank Japan, and CARDIoGRAMplusC4D Consortium for their publicly available summary data. The authors declare that no artificial intelligence program contributed to the compilation of the manuscript.

## Abbreviations

ABCG5/8: adenosine triphosphate-binding cassette transporters G5/8
ATP: adenosine triphosphate
CAD: coronary artery disease
eQTL: expression quantitative trait locus
FUMA: Functional Mapping and Annotation
GLGC: Global Lipids Genetics Consortium
GWAS: genome-wide association studies
ICD: International Classification of Diseases
IVW: inverse variance weighting
LDL: low-density lipoprotein
MI: myocardial infarction
MR: Mendelian randomization
PCSK9: proprotein convertase subtilisin/kexin type 9
RCT: randomized controlled trial

## Data availability

This study has been conducted using the UK Biobank Resource under Application number 98032. Summary-level data analyzed are available in the website http://csg.sph.umich.edu/willer/public/glgc-lipids2021/ for GLGC, https://www.finngen.fi/en/access_results for FinnGen, https://pheweb.jp/downloads for Biobank Japan, and http://www.cardiogramplusc4d.org/data-downloads/ for CARDIoGRAMplusC4D Consortium. The R code for data analysis is shared in the GitHub https://github.com/YANGGYEMMA/LDL_and_gallstones.

## Author contributions

GY and SB designed the study. GY undertook analyses with feedback from AMM, DG, CMS and SB. GY drafted the manuscript with critical feedback and revisions from AMM, DG, CMS and SB. GY is the guarantor of this work and takes responsibility for the integrity of the data analysis. All authors read and approved the final version of the manuscript.

## Perspectives

### Competence in medical knowledge

Genetic evidence supports that different plasma LDL-cholesterol lowering pathways have opposing effects on risk of gallstone disease. Genetic mimics of statins are associated with lower risk of gallstone disease.

### Translational outlook

Additional research is needed to elucidate the mechanisms underlying the opposing associations of different plasma LDL-cholesterol lowering pathways with gallstone disease, and to investigate the short-term effect of statins on gallstone disease.

## Reference

1. Ference BA, Ginsberg HN, Graham I et al. Low-density lipoproteins cause atherosclerotic cardiovascular disease. 1. Evidence from genetic, epidemiologic, and clinical studies. A consensus statement from the European Atherosclerosis Society Consensus Panel. European heart journal 2017;38:2459–2472.

2. Lammert F, Gurusamy K, Ko CW et al. Gallstones. Nature reviews Disease primers 2016;2:16024.

3. Nissen SE, Lincoff AM, Brennan D et al. Bempedoic Acid and Cardiovascular Outcomes in Statin-Intolerant Patients. The New England journal of medicine 2023;388:1353–1364.

4. Baigent C, Landray MJ, Reith C et al. The effects of lowering LDL cholesterol with simvastatin plus ezetimibe in patients with chronic kidney disease (Study of Heart and Renal Protection): a randomised placebo-controlled trial. Lancet (London, England) 2011;377:2181–92.

5. Bodmer M, Brauchli YB, Krähenbühl S, Jick SS, Meier CR. Statin use and risk of gallstone disease followed by cholecystectomy. Jama 2009;302:2001–7.

6. Erichsen R, Frøslev T, Lash TL, Pedersen L, Sørensen HT. Long-term statin use and the risk of gallstone disease: A population-based case-control study. American journal of epidemiology 2011;173:162–70.

7. Lawlor DA, Harbord RM, Sterne JA, Timpson N, Davey Smith G. Mendelian randomization: using genes as instruments for making causal inferences in epidemiology. Statistics in medicine 2008;27:1133–63.

8. Chen L, Yang H, Li H, He C, Yang L, Lv G. Insights into modifiable risk factors of cholelithiasis: A Mendelian randomization study. Hepatology (Baltimore, Md) 2022;75:785–796.

9. Stender S, Frikke-Schmidt R, Benn M, Nordestgaard BG, Tybjærg-Hansen A. Low-density lipoprotein cholesterol and risk of gallstone disease: a Mendelian randomization study and meta-analyses. Journal of hepatology 2013;58:126–33.

10. Luo J, Yang H, Song BL. Mechanisms and regulation of cholesterol homeostasis. Nature reviews Molecular cell biology 2020;21:225–245.

11. Ray KK, Corral P, Morales E, Nicholls SJ. Pharmacological lipid-modification therapies for prevention of ischaemic heart disease: current and future options. Lancet (London, England) 2019;394:697–708.

12. Ahmed O, Littmann K, Gustafsson U et al. Ezetimibe in Combination With Simvastatin Reduces Remnant Cholesterol Without Affecting Biliary Lipid Concentrations in Gallstone Patients. Journal of the American Heart Association 2018;7:e009876.

13. Stender S, Frikke-Schmidt R, Nordestgaard BG, Tybjærg-Hansen A. The ABCG5/8 cholesterol transporter and myocardial infarction versus gallstone disease. Journal of the American College of Cardiology 2014;63:2121–2128.

14. Bycroft C, Freeman C, Petkova D et al. The UK Biobank resource with deep phenotyping and genomic data. Nature 2018;562:203–209.

15. Kurki MI, Karjalainen J, Palta P et al. FinnGen provides genetic insights from a well-phenotyped isolated population. Nature 2023;613:508–518.

16. Sakaue S, Kanai M, Tanigawa Y et al. A cross-population atlas of genetic associations for 220 human phenotypes. Nature genetics 2021;53:1415–1424.

17. Giambartolomei C, Vukcevic D, Schadt EE et al. Bayesian test for colocalisation between pairs of genetic association studies using summary statistics. PLoS genetics 2014;10:e1004383.

18. Foley CN, Mason AM, Kirk PDW, Burgess S. MR-Clust: clustering of genetic variants in Mendelian randomization with similar causal estimates. Bioinformatics (Oxford, England) 2021;37:531–541.

19. Fairfield CJ, Drake TM, Pius R et al. Genome-wide analysis identifies gallstone-susceptibility loci including genes regulating gastrointestinal motility. Hepatology (Baltimore, Md) 2022;75:1081–1094.

20. Astle WJ, Elding H, Jiang T et al. The Allelic Landscape of Human Blood Cell Trait Variation and Links to Common Complex Disease. Cell 2016;167:1415–1429.e19.

21. Graham SE, Clarke SL, Wu KH et al. The power of genetic diversity in genome-wide association studies of lipids. Nature 2021;600:675–679.

22. Burgess S, Dudbridge F, Thompson SG. Combining information on multiple instrumental variables in Mendelian randomization: comparison of allele score and summarized data methods. Statistics in medicine 2016;35:1880–906.

23. Bowden J, Del Greco MF, Minelli C, Davey Smith G, Sheehan NA, Thompson JR. Assessing the suitability of summary data for two-sample Mendelian randomization analyses using MR-Egger regression: the role of the I2 statistic. International journal of epidemiology 2016;45:1961–1974.

24. Staley JR, Blackshaw J, Kamat MA et al. PhenoScanner: a database of human genotype-phenotype associations. Bioinformatics (Oxford, England) 2016;32:3207–3209.

25. Kamat MA, Blackshaw JA, Young R et al. PhenoScanner V2: an expanded tool for searching human genotype-phenotype associations. Bioinformatics (Oxford, England) 2019;35:4851–4853.

26. Nikpay M, Goel A, Won HH et al. A comprehensive 1,000 Genomes-based genome-wide association meta-analysis of coronary artery disease. Nature genetics 2015;47:1121–1130.

27. Burgess S, Butterworth A, Thompson SG. Mendelian randomization analysis with multiple genetic variants using summarized data. Genetic epidemiology 2013;37:658–65.

28. Bowden J, Davey Smith G, Haycock PC, Burgess S. Consistent Estimation in Mendelian Randomization with Some Invalid Instruments Using a Weighted Median Estimator. Genetic epidemiology 2016;40:304–14.

29. Bowden J, Davey Smith G, Burgess S. Mendelian randomization with invalid instruments: effect estimation and bias detection through Egger regression. International journal of epidemiology 2015;44:512–25.

30. Paternoster R, Brame R, Mazerolle P, Piquero A. Using the Correct Statistical Test for Equality of Regression Coefficients. Criminology 1998;36:859–866.

31. Zuber V, Grinberg NF, Gill D et al. Combining evidence from Mendelian randomization and colocalization: Review and comparison of approaches. American journal of human genetics 2022;109:767–782.

32. Watanabe K, Taskesen E, van Bochoven A, Posthuma D. Functional mapping and annotation of genetic associations with FUMA. Nature communications 2017;8:1826.

33. Preiss D, Tikkanen MJ, Welsh P et al. Lipid-modifying therapies and risk of pancreatitis: a meta-analysis. Jama 2012;308:804–11.

34. Tazuma S, Kajiyama G, Mizuno T et al. A combination therapy with simvastatin and ursodeoxycholic acid is more effective for cholesterol gallstone dissolution than is ursodeoxycholic acid monotherapy. Journal of clinical gastroenterology 1998;26:287–91.

35. Logan GM, Duane WC. Lovastatin added to ursodeoxycholic acid further reduces biliary cholesterol saturation. Gastroenterology 1990;98:1572–6.

36. Binnington B, Nguyen L, Kamani M et al. Inhibition of Rab prenylation by statins induces cellular glycosphingolipid remodeling. Glycobiology 2016;26:166–80.

37. Lee BJ, Kim JS, Kim BK et al. Effects of sphingolipid synthesis inhibition on cholesterol gallstone formation in C57BL/6J mice. Journal of gastroenterology and hepatology 2010;25:1105–10.

38. Yang G, Schooling CM. Statins, Type 2 Diabetes, and Body Mass Index: A Univariable and Multivariable Mendelian Randomization Study. The Journal of clinical endocrinology and metabolism 2023;108:385–396.

39. Lotta LA, Sharp SJ, Burgess S et al. Association Between Low-Density Lipoprotein Cholesterol-Lowering Genetic Variants and Risk of Type 2 Diabetes: A Meta-analysis. Jama 2016;316:1383–1391.

40. Shaffer EA. Gallstone disease: Epidemiology of gallbladder stone disease. Best practice & research Clinical gastroenterology 2006;20:981–96.

41. Schooling CM, Zhao JV, Au Yeung SL, Leung GM. Investigating pleiotropic effects of statins on ischemic heart disease in the UK Biobank using Mendelian randomisation. eLife 2020;9.

42. Li S, Schooling CM. A phenome-wide association study of genetically mimicked statins. BMC medicine 2021;19:151.

