## Supplemental Figures 1-9, Supplemental Tables 1-9 for "Trans-biobank Mendelian randomization analyses identify opposing pathways in plasma low-density lipoprotein-cholesterol lowering and gallstone disease"

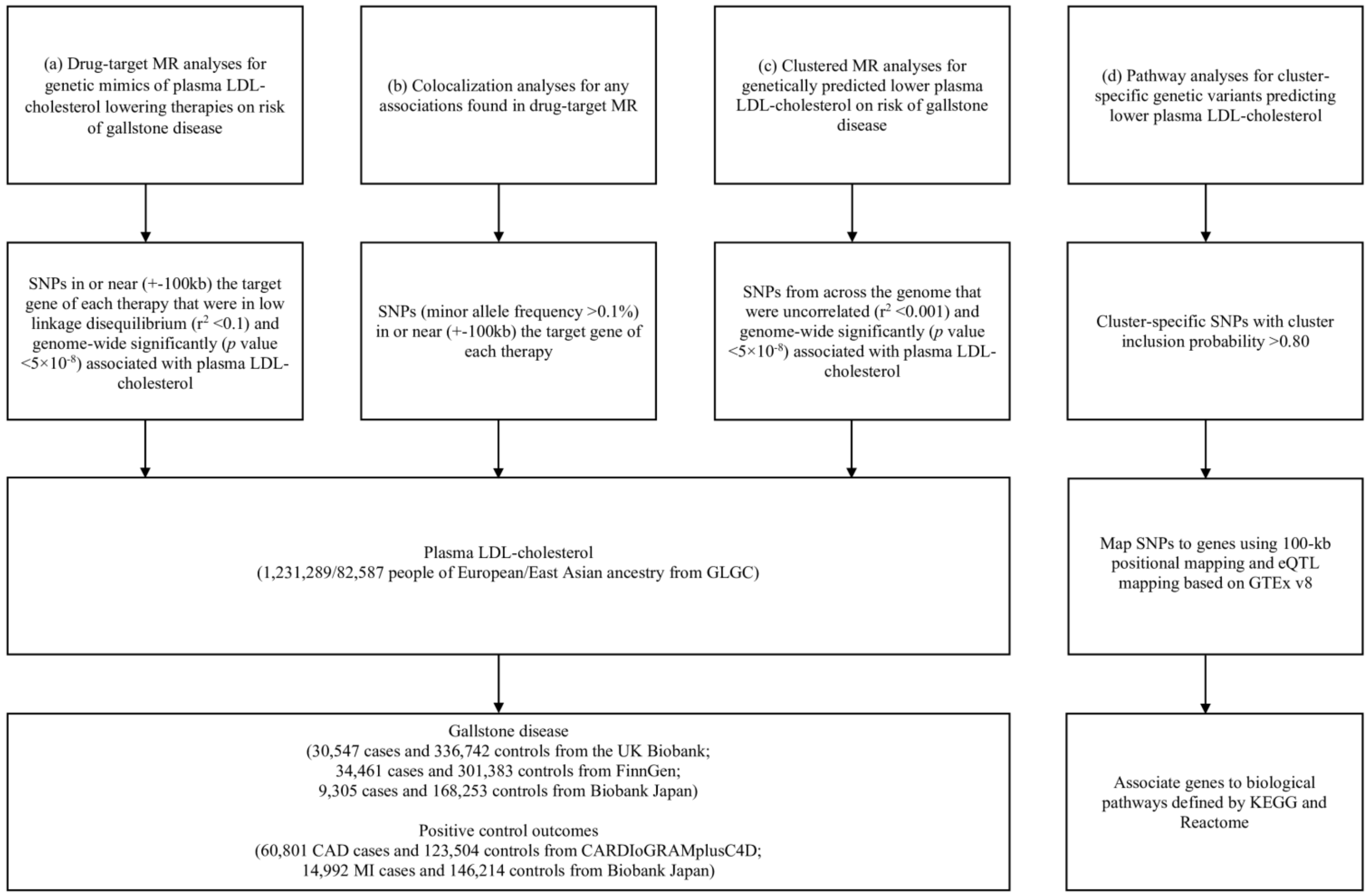


Supplemental Figure 1. Flowchart of the study design.

CAD, coronary artery disease; CARDIoGRAMplusC4D, Coronary ARtery DIsease Genome wide Replication and Meta-analysis plus The Coronary Artery Disease Genetics; eQTL, expression quantitative trait locus; GLGC, Global Lipids Genetics Consortium; LDL, low-density lipoprotein; MI, myocardial infarction; MR, Mendelian randomization.

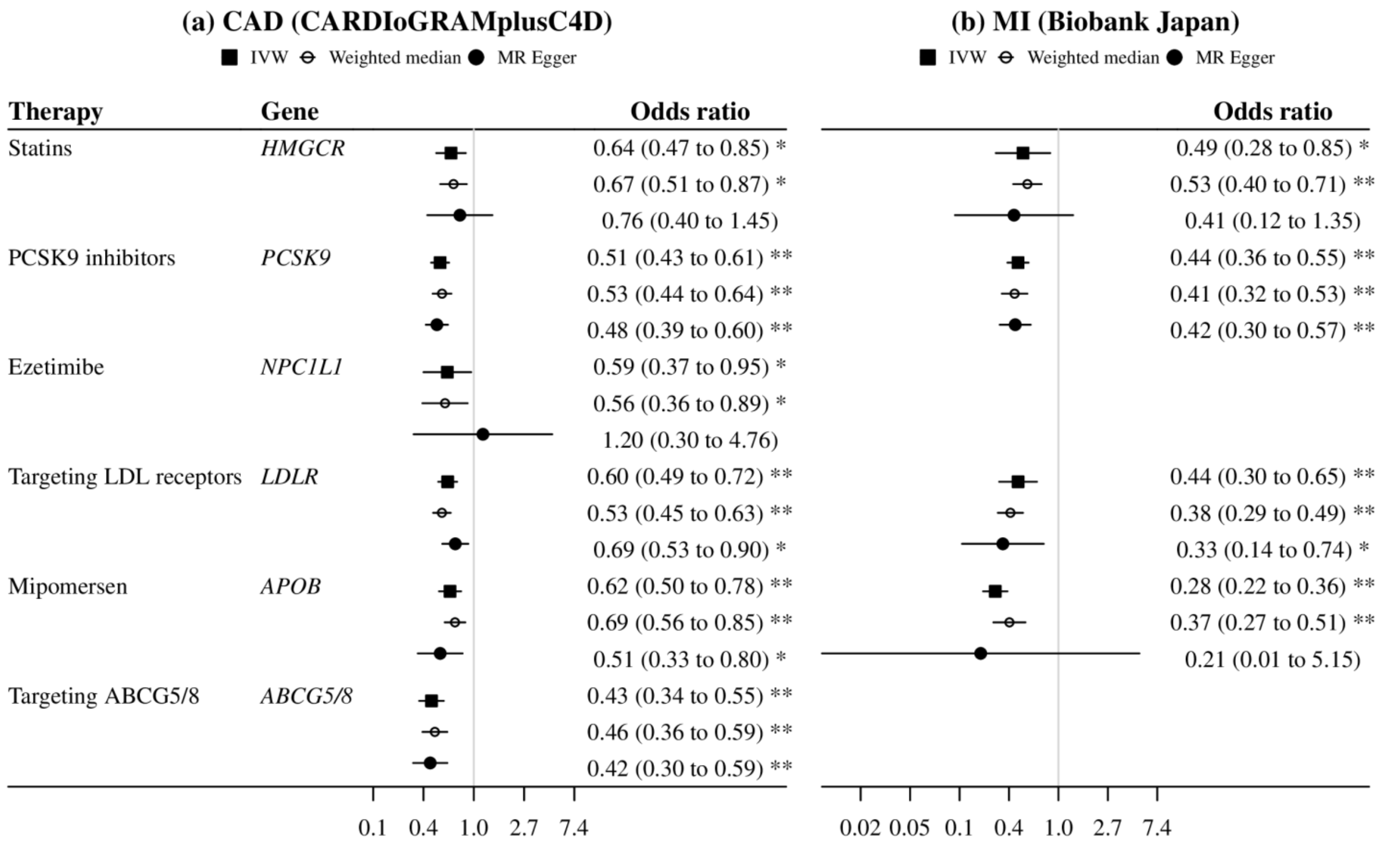


Supplemental Figure 2. Mendelian randomization estimates for genetic mimics of plasma LDL-cholesterol lowering therapies on risk of CAD and MI. ABCG5/8, adenosine triphosphate (ATP)-binding cassette transporters G5/8; CAD, coronary artery disease; CARDIoGRAMplusC4D, Coronary ARtery DIsease Genome wide Replication and Meta-analysis plus The Coronary Artery Disease Genetics; IVW, inverse variance weighted; LDL, low-density lipoprotein; MI, myocardial infarction; PCSK9, proprotein convertase subtilisin/kexin type 9. Estimates are expressed in odds ratio per 1-standard deviation (around 0.87 mmol/L) reduction in plasma LDL-cholesterol. * denotes *p* value <0.05; ** denotes *p* value <0.001.

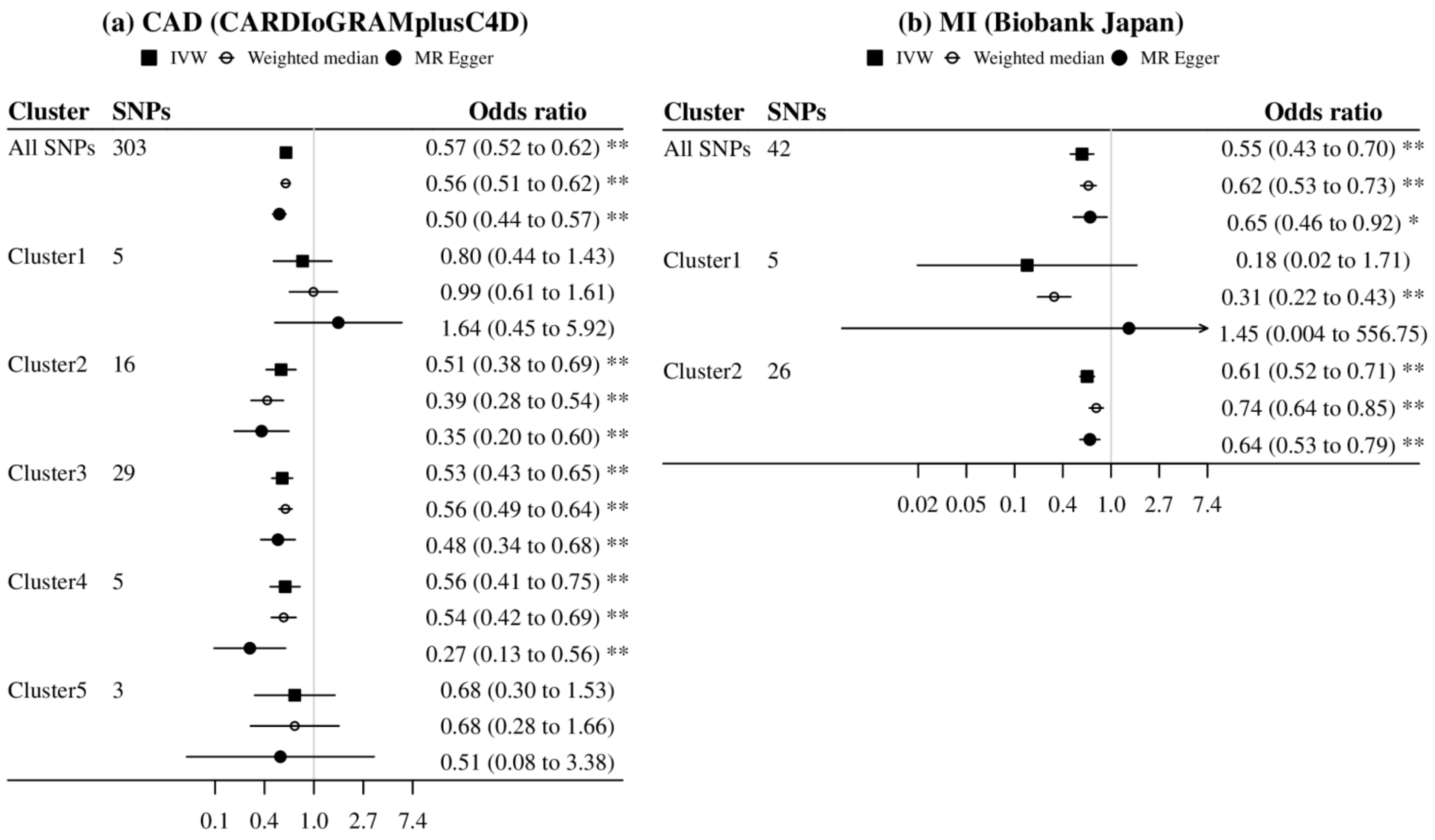


Supplemental Figure 3. Mendelian randomization estimates for genetically predicted lower LDL-cholesterol on risk of CAD and MI using all SNPs and cluster-specific SNPs (inclusion probability >0.80).

CAD, coronary artery disease; CARDIoGRAMplusC4D, Coronary ARtery DIsease Genome wide Replication and Meta-analysis plus The Coronary Artery Disease Genetics; IVW, inverse variance weighted; LDL, low-density lipoprotein; MI, myocardial infarction. Estimates are expressed in odds ratio per 1-standard deviation (around 0.87 mmol/L) reduction in plasma LDL-cholesterol. * denotes $p$ value <0.05; ** denotes $p$ value <0.001.

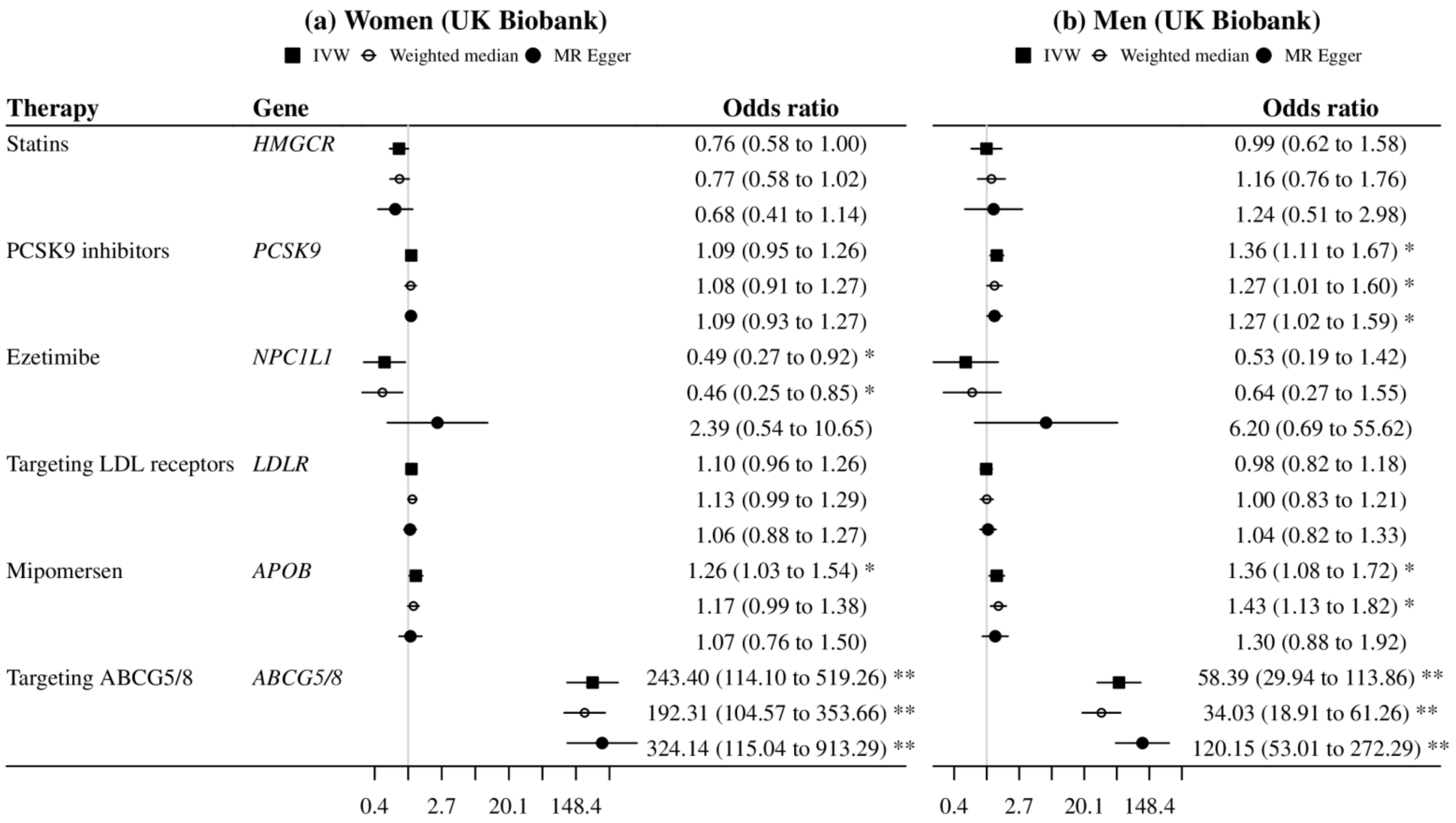


Supplemental Figure 4. Sex-specific Mendelian randomization estimates for genetic mimics of plasma LDL-cholesterol lowering therapies on risk of gallstone disease in the UK Biobank.

ABCG5/8, adenosine triphosphate (ATP)-binding cassette transporters G5/8; IVW, inverse variance weighted; LDL, low-density lipoprotein; PCSK9, proprotein convertase subtilisin/kexin type 9. Estimates are expressed in odds ratio per 1-standard deviation (around 0.87 mmol/L) reduction in plasma LDL-cholesterol. * denotes *p* value <0.05; ** denotes *p* value <0.001.

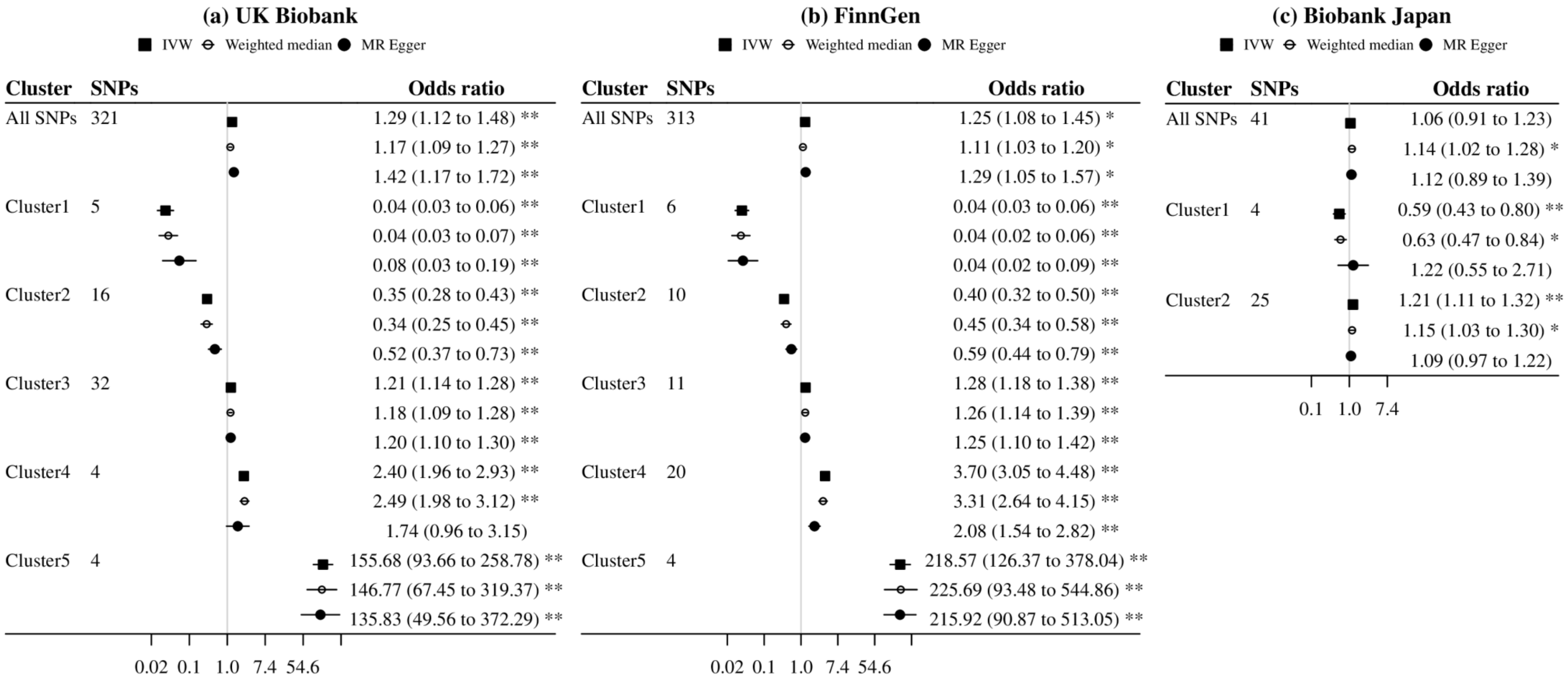


Supplemental Figure 5. Mendelian randomization estimates for genetically predicted lower LDL-cholesterol on risk of gallstone disease using all SNPs and cluster-specific SNPs (inclusion probability >0.80) in sensitivity analysis excluding SNPs associated with common confounders.

IVW, inverse variance weighted; LDL, low-density lipoprotein. The SNPs excluded for UK Biobank and FinnGen were rs11499828, rs174547 and rs3184504; the SNP excluded for Biobank Japan was rs4646776. Estimates are expressed in odds ratio per 1-standard deviation (around 0.87 mmol/L) reduction in plasma LDL-cholesterol. * denotes *p* value <0.05; ** denotes *p* value <0.001.

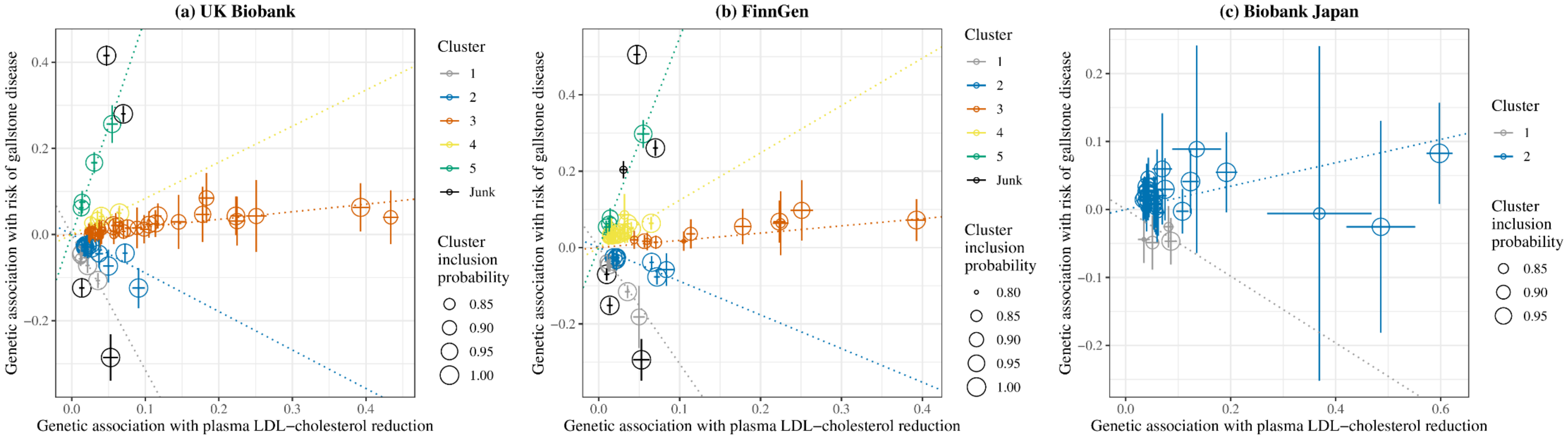


Supplemental Figure 6. Genetic associations with plasma LDL-cholesterol reduction (standard deviation) and risk of gallstone disease (log odds) for SNPs with inclusion probability >0.80 in clustered Mendelian randomization analyses excluding SNPs associated with common confounders.

LDL, low-density lipoprotein. The SNPs excluded for UK Biobank and FinnGen were rs11499828, rs174547 and rs3184504; the SNP excluded for Biobank Japan was rs4646776. Points represent SNPs; dotted lines are cluster means; error bars are 95% confidence intervals for genetic associations.

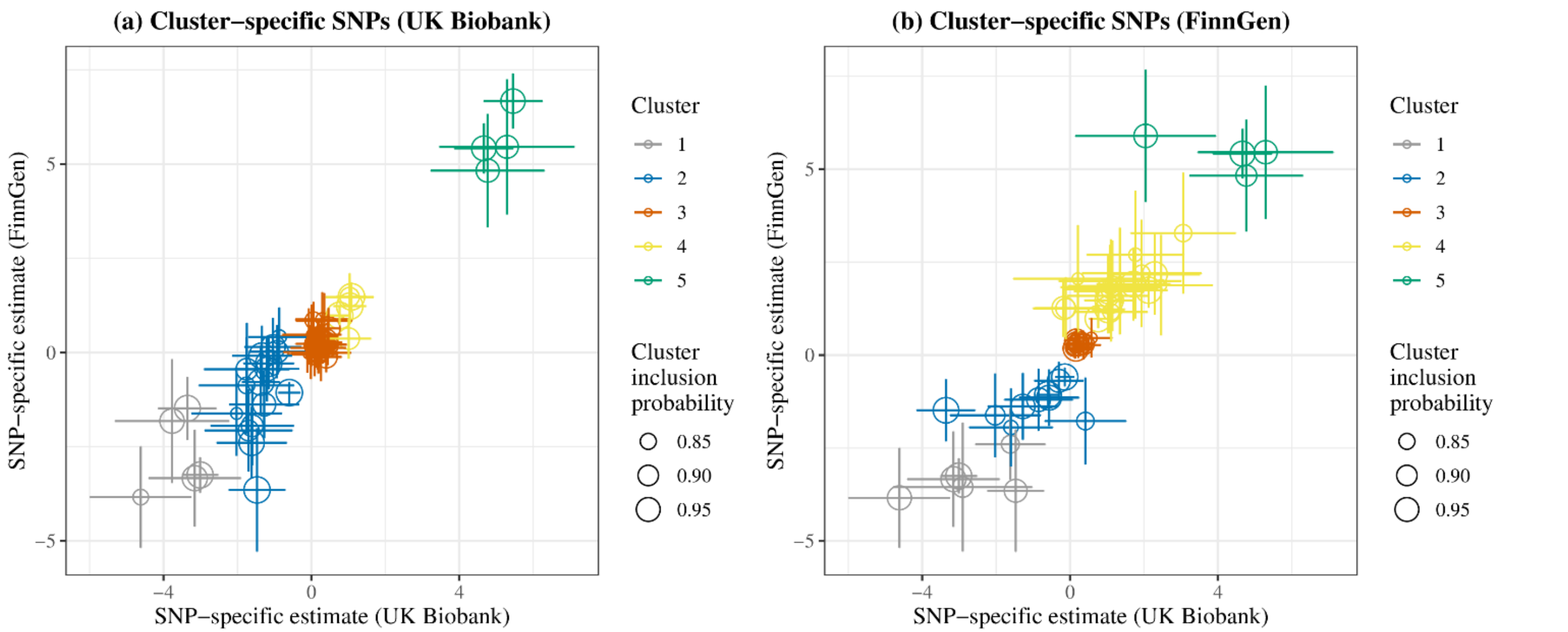


Supplemental Figure 7. SNP-specific estimates for genetically predicted lower plasma LDL-cholesterol on risk of gallstone disease in the UK Biobank and FinnGen for SNPs with inclusion probability >0.80 in clustered Mendelian randomization analyses.

LDL, low-density lipoprotein. Points represent SNPs; error bars are 95% confidence intervals for SNP-specific estimates. Estimates are expressed in log odds per 1-standard deviation (around 0.87 mmol/L) reduction in plasma LDL-cholesterol.

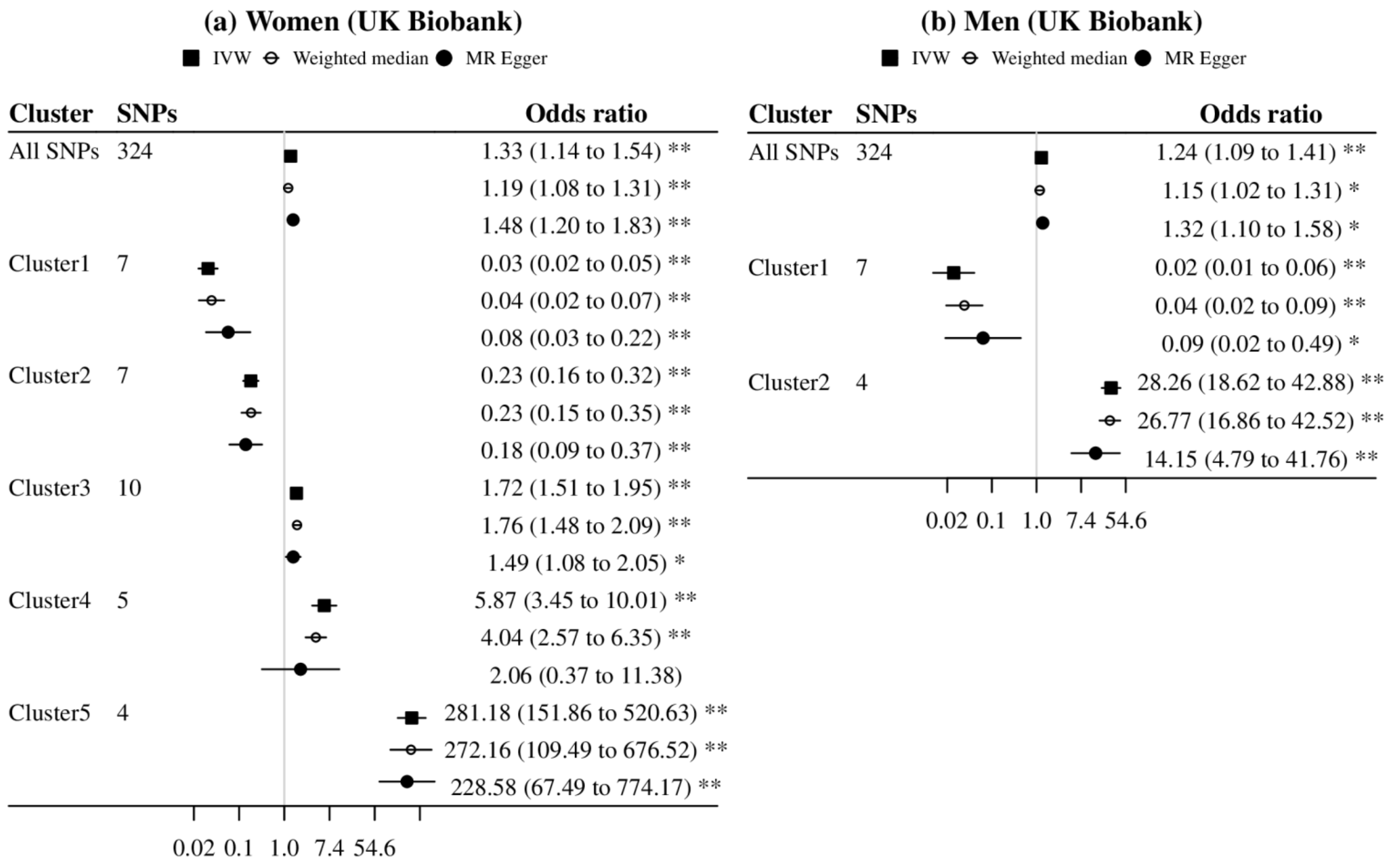


Supplemental Figure 8. Sex-specific Mendelian randomization estimates for genetically predicted lower LDL-cholesterol on risk of gallstone disease using all SNPs and cluster-specific SNPs (inclusion probability >0.80).

IVW, inverse variance weighted; LDL, low-density lipoprotein. Estimates are expressed in odds ratio per 1-standard deviation (around 0.87 mmol/L) reduction in plasma LDL-cholesterol. * denotes *p* value <0.05; ** denotes *p* value <0.001.

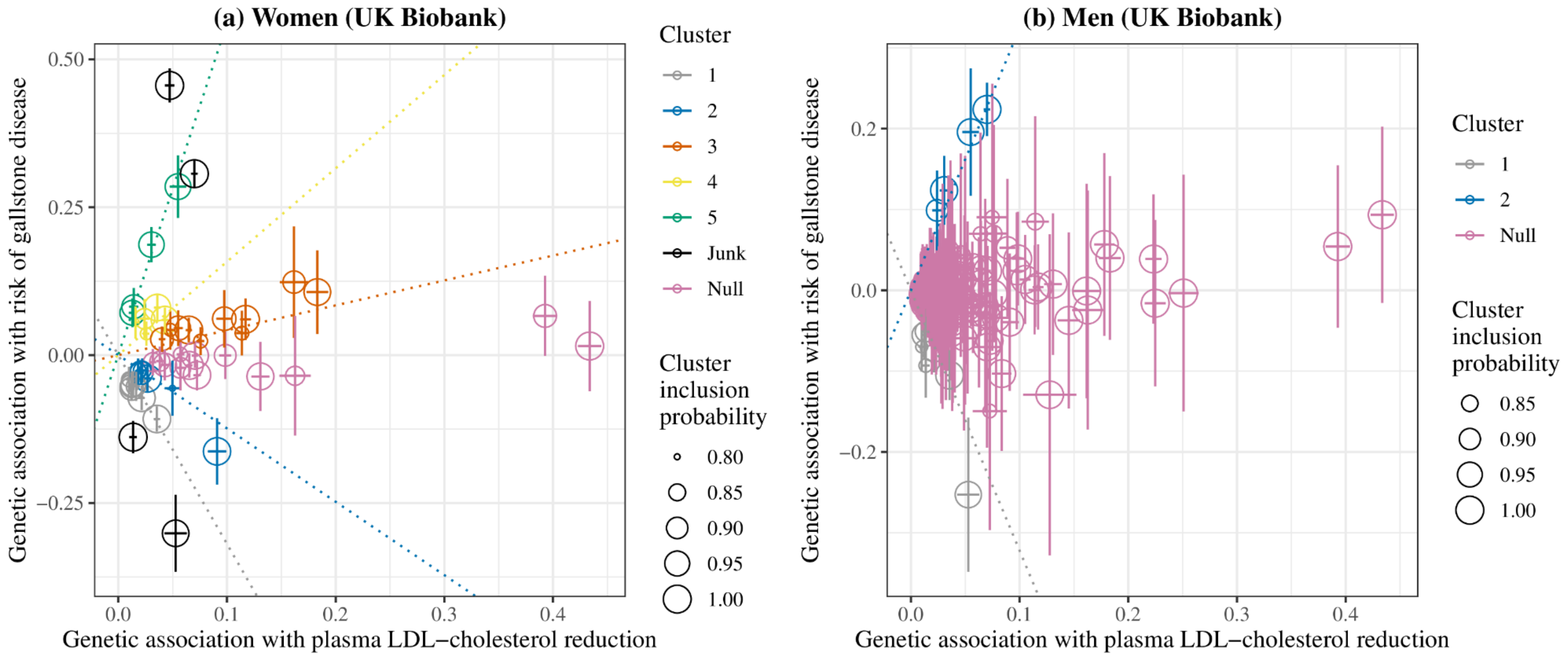


Supplemental Figure 9. Sex-specific genetic associations with plasma LDL-cholesterol reduction (standard deviation) and risk of gallstone disease (log odds) for SNPs with inclusion probability >0.80 in clustered Mendelian randomization analyses in the UK Biobank.

LDL, low-density lipoprotein. Points represent SNPs; dotted lines are cluster means; error bars are 95% confidence intervals for genetic associations.

Supplemental Table 1. Definition codes of gallstone disease across the three biobanks.

| Biobank | Definition | ICD-10 | ICD-9 | ICD-8 | UKB | OPCS4 | OPCS3 |
|---|---|---|---|---|---|---|---|
| UK Biobank | Gallstone-related pathology | K56.3, K80, K81, K85.1, K91.5, K91.86 | 560.31, 574, 575.0, 575.1, 576.0, 997.41 | - | 1160-1163 | - | - |
| | Gallstone-related procedures | - | - | - | 1455, 1528 | J18, J21.1, J24.2, J24.3, J26.1, J33.1, J33.2, J38.1, J41.1, J41.3, J42.3, J49.1, J49.2, J52.1, J60.2, J68.1, J76.1 | 511, 521, 522, 530.2 |
| | Alternative pathology | C17.0, C22-25, D01.5, D13.2, D13.4-13.7, S36.1, K82.4, K83.5 | 152.0, 155-157, 230.8, 211.2, 211.5-211.7, 230.8, 575.6, 864, 868.02, 868.12 | - | 1024-1026 | - | - |
| FinnGen | Cholelithiasis, broad definition with cholecystitis | K80, K81 | 574, 575.0, 575.1 | 574, 5750.0-5750.3 | - | - | - |
| Biobank Japan | Cholelithiasis | K80 | - | - | - | - | - |

ICD, International Classification of Diseases; OPCS, Office of Population Censuses and Surveys Classification of Surgical Operations and Procedures; UKB, UK Biobank codes for self-reported conditions verified by medical notes. UK Biobank participants with codes for gallstone-related pathology or procedures were defined as cases, but those who underwent gallstone-related procedures with alternative pathology were excluded from the analysis.

Supplemental Table 2. Description of GWAS data used in this study.

| Phenotype | Study | Ancestry | Sample size | Covariates |
|---|---|---|---|---|
| Gallstone disease | UK Biobank | European | Cases = 21,201 women/9,346 men<br>Controls = 177,478 women/159,264 men | Age, age^2, sex, age * sex, age^2 * sex, and the first 20 PCs in sex-combined analyses; age, age^2 and the first 20 PCs in sex-specific analyses |
| Gallstone disease | FinnGen | European | Cases = 34,461<br>Controls = 301,383 | Age, sex, genotyping batch, and ten PCs |
| Gallstone disease | Biobank Japan | East Asian | Cases = 9,305<br>Controls = 168,253 | Age, age^2, sex, age * sex, age^2 * sex, and the first 20 PCs |
| Plasma LDL-cholesterol | GLGC | European | *N* = 1,231,289 | Age, age^2, sex, PCs of ancestry and study-specific covariates |
| Plasma LDL-cholesterol | GLGC | East Asian | *N* = 82,587 | Age, age^2, sex, PCs of ancestry and study-specific covariates |
| Coronary artery disease | CARDIoGRAM plusC4D | Mixed (mainly European) | Cases = 60,801<br>Controls = 123,504 | Study-specific covariates |
| Myocardial infarction | Biobank Japan | East Asian | Cases = 14,992<br>Controls = 146,214 | Age, age^2, sex, age * sex, age^2 * sex, and the first 20 PCs |

CARDIoGRAMplusC4D, Coronary ARtery DIsease Genome wide Replication and Meta-analysis plus The Coronary Artery Disease Genetics; GLGC, Global Lipids Genetics Consortium; GWAS, genome-wide association studies; LDL, low-density lipoprotein; PC, principal components.

Supplemental Table 3. SNP-specific estimates for genetic mimics of plasma LDL-cholesterol lowering therapies on plasma LDL-cholesterol (standard deviation) in people of European and East Asian ancestry.

| Ancestry | Therapy | Gene | SNP | Effect allele | Other allele | Effect allele frequency | Beta | SE | *P* value | F-statistic |
|---|---|---|---|---|---|---|---|---|---|---|
| European | Statins | *HMGCR* | rs375392181 | T | G | 0.010 | 0.0454 | 0.0071 | 3.0E-08 | 41.3 |
| European | Statins | *HMGCR* | rs72768351 | G | A | 0.019 | -0.0401 | 0.0055 | 4.4E-10 | 52.3 |
| European | Statins | *HMGCR* | rs75144964 | G | C | 0.016 | -0.0403 | 0.0060 | 6.4E-09 | 45.3 |
| European | Statins | *HMGCR* | rs1423527 | A | C | 0.399 | 0.0653 | 0.0014 | <4.9E-324 | 2127.3 |
| European | Statins | *HMGCR* | rs35122945 | C | A | 0.067 | -0.0371 | 0.0030 | 5.2E-27 | 155.6 |
| European | Statins | *HMGCR* | rs115664150 | T | C | 0.062 | 0.0431 | 0.0029 | 9.9E-38 | 221.4 |
| European | Statins | *HMGCR* | rs75240579 | T | C | 0.040 | -0.0404 | 0.0038 | 1.7E-20 | 115.7 |
| European | Statins | *HMGCR* | rs112672253 | T | A | 0.008 | 0.0576 | 0.0083 | 1.9E-09 | 48.4 |
| European | Statins | *HMGCR* | rs17244939 | C | A | 0.016 | -0.0430 | 0.0062 | 1.7E-09 | 48.8 |
| European | Statins | *HMGCR* | rs115169875 | A | G | 0.026 | -0.0337 | 0.0046 | 2.9E-10 | 53.4 |
| European | Statins | *HMGCR* | rs151264833 | G | C | 0.024 | 0.0340 | 0.0046 | 1.5E-10 | 55.0 |
| European | Statins | *HMGCR* | rs62366588 | A | C | 0.058 | -0.0346 | 0.0032 | 1.0E-20 | 117.0 |
| European | Statins | *HMGCR* | rs74695562 | G | T | 0.042 | -0.0369 | 0.0037 | 7.1E-18 | 99.7 |
| European | Statins | *HMGCR* | rs78587954 | G | C | 0.065 | 0.0526 | 0.0028 | 2.7E-58 | 348.1 |
| European | Statins | *HMGCR* | rs144083983 | T | C | 0.070 | -0.0418 | 0.0030 | 1.9E-33 | 195.2 |
| European | Statins | *HMGCR* | rs181668591 | T | C | 0.010 | 0.0604 | 0.0072 | 5.6E-13 | 69.8 |
| European | Statins | *HMGCR* | rs114253542 | C | T | 0.007 | 0.0759 | 0.0087 | 3.9E-14 | 76.8 |
| European | Statins | *HMGCR* | rs180755046 | A | C | 0.016 | -0.0421 | 0.0062 | 5.5E-09 | 45.7 |
| European | Statins | *HMGCR* | rs182826525 | G | A | 0.014 | 0.1035 | 0.0086 | 4.2E-25 | 143.9 |

| | | | | | | | | | | |
|---|---|---|---|---|---|---|---|---|---|---|
| European | Statins | *HMGCR* | rs151000110 | A | G | 0.057 | 0.0670 | 0.0031 | 1.5E-77 | 466.7 |
| European | Statins | *HMGCR* | rs200823803 | C | T | 0.018 | 0.0551 | 0.0060 | 2.2E-15 | 84.5 |
| European | PCSK9 inhibitors | *PCSK9* | rs12043403 | C | T | 0.098 | -0.0245 | 0.0025 | 1.0E-17 | 98.7 |
| European | PCSK9 inhibitors | *PCSK9* | rs12409233 | C | G | 0.088 | -0.0226 | 0.0024 | 1.6E-15 | 85.3 |
| European | PCSK9 inhibitors | *PCSK9* | rs11206498 | G | A | 0.342 | 0.0103 | 0.0015 | 1.4E-09 | 49.3 |
| European | PCSK9 inhibitors | *PCSK9* | rs890576 | G | C | 0.202 | -0.0153 | 0.0017 | 4.6E-14 | 76.4 |
| European | PCSK9 inhibitors | *PCSK9* | rs145075626 | G | C | 0.017 | -0.0503 | 0.0055 | 3.0E-15 | 83.6 |
| European | PCSK9 inhibitors | *PCSK9* | rs17111474 | T | C | 0.296 | 0.0113 | 0.0016 | 2.4E-09 | 47.9 |
| European | PCSK9 inhibitors | *PCSK9* | rs77875082 | A | G | 0.031 | 0.0538 | 0.0042 | 3.4E-28 | 162.8 |
| European | PCSK9 inhibitors | *PCSK9* | rs2495503 | A | G | 0.258 | 0.0349 | 0.0016 | 2.3E-80 | 484.1 |
| European | PCSK9 inhibitors | *PCSK9* | rs34232196 | T | C | 0.242 | -0.0663 | 0.0016 | 3.0E-266 | 1632.3 |
| European | PCSK9 inhibitors | *PCSK9* | rs374459115 | A | G | 0.017 | 0.0683 | 0.0068 | 2.8E-18 | 102.2 |
| European | PCSK9 inhibitors | *PCSK9* | rs28775984 | C | T | 0.432 | -0.0193 | 0.0030 | 4.3E-08 | 40.3 |
| European | PCSK9 inhibitors | *PCSK9* | rs182491400 | T | C | 0.021 | 0.0445 | 0.0056 | 4.9E-12 | 64.1 |
| European | PCSK9 inhibitors | *PCSK9* | rs181331606 | G | C | 0.090 | 0.0286 | 0.0030 | 1.0E-16 | 92.6 |
| European | PCSK9 inhibitors | *PCSK9* | rs12739979 | T | C | 0.238 | -0.0270 | 0.0018 | 6.3E-38 | 222.6 |
| European | PCSK9 inhibitors | *PCSK9* | rs11810371 | A | G | 0.043 | -0.0423 | 0.0037 | 2.7E-23 | 132.8 |
| European | PCSK9 inhibitors | *PCSK9* | rs72660548 | G | C | 0.020 | 0.0704 | 0.0050 | 1.2E-34 | 202.4 |
| European | PCSK9 inhibitors | *PCSK9* | rs17111503 | G | A | 0.256 | 0.0487 | 0.0016 | 6.8E-151 | 919.5 |
| European | PCSK9 inhibitors | *PCSK9* | rs2479408 | G | C | 0.199 | -0.0445 | 0.0018 | 1.7E-100 | 608.3 |
| European | PCSK9 inhibitors | *PCSK9* | rs11591147 | T | G | 0.016 | -0.4337 | 0.0054 | <4.9E-324 | 6388.7 |
| European | PCSK9 inhibitors | *PCSK9* | rs479832 | C | T | 0.178 | -0.0144 | 0.0019 | 1.8E-11 | 60.7 |
| European | PCSK9 inhibitors | *PCSK9* | rs41294825 | T | A | 0.043 | -0.0414 | 0.0034 | 1.2E-25 | 147.2 |

| | | | | | | | | | | |
|---|---|---|---|---|---|---|---|---|---|---|
| European | PCSK9 inhibitors | *PCSK9* | rs693668 | G | A | 0.359 | -0.0565 | 0.0015 | 2.6E-243 | 1490.6 |
| European | PCSK9 inhibitors | *PCSK9* | rs7525503 | T | G | 0.023 | 0.0713 | 0.0050 | 6.2E-35 | 204.2 |
| European | PCSK9 inhibitors | *PCSK9* | rs11206517 | G | T | 0.037 | 0.0889 | 0.0037 | 7.6E-95 | 573.4 |
| European | PCSK9 inhibitors | *PCSK9* | rs41297885 | G | C | 0.036 | -0.0406 | 0.0038 | 2.2E-20 | 115.0 |
| European | PCSK9 inhibitors | *PCSK9* | rs28385715 | G | T | 0.023 | 0.0456 | 0.0049 | 9.9E-16 | 86.6 |
| European | PCSK9 inhibitors | *PCSK9* | rs77011887 | T | C | 0.017 | 0.0459 | 0.0057 | 3.7E-12 | 64.8 |
| European | PCSK9 inhibitors | *PCSK9* | rs142116310 | A | G | 0.011 | 0.0554 | 0.0072 | 3.8E-11 | 58.7 |
| European | PCSK9 inhibitors | *PCSK9* | rs115465289 | A | G | 0.031 | -0.0407 | 0.0042 | 1.0E-16 | 92.7 |
| European | PCSK9 inhibitors | *PCSK9* | rs12031153 | A | G | 0.053 | 0.0331 | 0.0035 | 1.5E-16 | 91.6 |
| European | PCSK9 inhibitors | *PCSK9* | rs138779133 | G | A | 0.027 | -0.0599 | 0.0043 | 9.1E-33 | 190.9 |
| European | PCSK9 inhibitors | *PCSK9* | rs79494709 | C | G | 0.049 | -0.0258 | 0.0033 | 8.3E-12 | 62.7 |
| European | PCSK9 inhibitors | *PCSK9* | rs55817205 | A | G | 0.012 | 0.0676 | 0.0071 | 2.6E-16 | 90.1 |
| European | Ezetimibe | *NPC1L1* | rs77826622 | C | T | 0.022 | 0.0313 | 0.0049 | 4.4E-08 | 40.2 |
| European | Ezetimibe | *NPC1L1* | rs17725246 | C | T | 0.195 | 0.0428 | 0.0018 | 1.3E-94 | 572.0 |
| European | Ezetimibe | *NPC1L1* | rs77517259 | T | C | 0.010 | 0.0503 | 0.0073 | 2.6E-09 | 47.6 |
| European | Ezetimibe | *NPC1L1* | rs217370 | G | A | 0.467 | -0.0254 | 0.0014 | 1.3E-55 | 331.6 |
| European | Ezetimibe | *NPC1L1* | rs79854399 | T | C | 0.019 | -0.0390 | 0.0052 | 1.2E-10 | 55.8 |
| European | Ezetimibe | *NPC1L1* | rs143116067 | A | G | 0.068 | -0.0201 | 0.0029 | 3.7E-09 | 46.7 |
| European | Targeting LDL receptors | *LDLR* | rs148724334 | C | G | 0.017 | 0.0428 | 0.0058 | 2.6E-10 | 53.7 |
| European | Targeting LDL receptors | *LDLR* | rs117210556 | C | T | 0.065 | 0.0371 | 0.0028 | 5.8E-30 | 173.7 |
| European | Targeting LDL receptors | *LDLR* | rs118115977 | T | C | 0.014 | 0.0394 | 0.0062 | 4.2E-08 | 40.4 |
| European | Targeting LDL receptors | *LDLR* | rs12609589 | T | C | 0.157 | 0.0445 | 0.0020 | 3.4E-82 | 495.4 |
| European | Targeting LDL receptors | *LDLR* | rs73013176 | C | T | 0.012 | -0.2141 | 0.0068 | 2.4E-164 | 1002.6 |

| | | | | | | | | | | |
|---|---|---|---|---|---|---|---|---|---|---|
| European | Targeting LDL receptors | *LDLR* | rs10420325 | A | T | 0.350 | 0.0157 | 0.0023 | 2.0E-09 | 48.3 |
| European | Targeting LDL receptors | *LDLR* | rs138736573 | G | A | 0.009 | 0.0692 | 0.0074 | 1.1E-15 | 86.3 |
| European | Targeting LDL receptors | *LDLR* | rs36049922 | C | T | 0.035 | 0.0437 | 0.0040 | 1.7E-21 | 121.9 |
| European | Targeting LDL receptors | *LDLR* | rs56315738 | T | C | 0.008 | -0.1117 | 0.0089 | 2.0E-27 | 158.2 |
| European | Targeting LDL receptors | *LDLR* | rs73015007 | A | G | 0.241 | -0.0757 | 0.0016 | <4.9E-324 | 2127.3 |
| European | Targeting LDL receptors | *LDLR* | rs10423733 | C | T | 0.171 | -0.1315 | 0.0019 | <4.9E-324 | 4977.0 |
| European | Targeting LDL receptors | *LDLR* | rs112159161 | T | C | 0.015 | -0.2000 | 0.0059 | 1.6E-187 | 1145.9 |
| European | Targeting LDL receptors | *LDLR* | rs146335137 | T | C | 0.009 | 0.0618 | 0.0077 | 5.6E-12 | 63.8 |
| European | Targeting LDL receptors | *LDLR* | rs17242353 | T | C | 0.032 | 0.1091 | 0.0043 | 5.9E-108 | 654.4 |
| European | Targeting LDL receptors | *LDLR* | rs17242367 | T | C | 0.069 | 0.0283 | 0.0029 | 3.9E-17 | 95.1 |
| European | Targeting LDL receptors | *LDLR* | rs17248748 | T | C | 0.018 | -0.0749 | 0.0056 | 5.5E-31 | 180.0 |
| European | Targeting LDL receptors | *LDLR* | rs6511721 | G | A | 0.483 | 0.0590 | 0.0014 | 4.3E-270 | 1656.1 |
| European | Targeting LDL receptors | *LDLR* | rs73015030 | A | G | 0.032 | -0.1334 | 0.0040 | 1.1E-179 | 1097.5 |
| European | Targeting LDL receptors | *LDLR* | rs560115066 | G | T | 0.036 | 0.0725 | 0.0045 | 6.6E-44 | 259.4 |
| European | Targeting LDL receptors | *LDLR* | rs62129100 | G | A | 0.172 | 0.0214 | 0.0032 | 8.4E-09 | 44.6 |
| European | Targeting LDL receptors | *LDLR* | rs147223423 | T | A | 0.010 | 0.0999 | 0.0071 | 5.7E-34 | 198.3 |
| European | Targeting LDL receptors | *LDLR* | rs17249001 | A | G | 0.071 | 0.0715 | 0.0029 | 2.3E-99 | 601.4 |
| European | Targeting LDL receptors | *LDLR* | rs3826810 | A | G | 0.043 | 0.0376 | 0.0034 | 2.9E-21 | 120.3 |
| European | Targeting LDL receptors | *LDLR* | rs5742911 | G | A | 0.312 | -0.0659 | 0.0015 | 6.0E-306 | 1877.7 |
| European | Targeting LDL receptors | *LDLR* | rs143587805 | T | A | 0.012 | 0.0631 | 0.0067 | 4.0E-16 | 89.0 |
| European | Targeting LDL receptors | *LDLR* | rs147540853 | A | G | 0.026 | -0.0911 | 0.0043 | 5.6E-75 | 450.9 |
| European | Targeting LDL receptors | *LDLR* | rs111622889 | T | C | 0.021 | 0.0447 | 0.0056 | 6.7E-12 | 63.3 |
| European | Targeting LDL receptors | *LDLR* | rs148362484 | T | C | 0.028 | 0.0286 | 0.0044 | 1.7E-08 | 42.8 |

| | | | | | | | | | | |
|---|---|---|---|---|---|---|---|---|---|---|
| European | Targeting LDL receptors | *LDLR* | rs189143685 | T | C | 0.011 | 0.0494 | 0.0069 | 6.1E-10 | 51.4 |
| European | Targeting LDL receptors | *LDLR* | rs72983207 | T | A | 0.018 | -0.0443 | 0.0056 | 1.1E-11 | 62.0 |
| European | Targeting LDL receptors | *LDLR* | rs4804573 | A | G | 0.467 | -0.0630 | 0.0014 | 1.9e-310 | 1905.5 |
| European | Targeting LDL receptors | *LDLR* | rs117339792 | A | G | 0.016 | -0.0846 | 0.0061 | 1.2E-32 | 190.1 |
| European | Targeting LDL receptors | *LDLR* | rs146576912 | T | C | 0.051 | -0.1555 | 0.0035 | <4.9E-324 | 1992.1 |
| European | Targeting LDL receptors | *LDLR* | rs4804149 | C | T | 0.288 | 0.0241 | 0.0016 | 1.8E-37 | 219.9 |
| European | Targeting LDL receptors | *LDLR* | rs379309 | C | T | 0.495 | 0.0252 | 0.0014 | 2.8E-52 | 311.0 |
| European | Targeting LDL receptors | *LDLR* | rs139198665 | T | C | 0.026 | 0.0614 | 0.0048 | 2.5E-28 | 163.6 |
| European | Targeting LDL receptors | *LDLR* | rs140842514 | C | A | 0.029 | -0.0283 | 0.0043 | 1.3E-08 | 43.4 |
| European | Targeting LDL receptors | *LDLR* | rs12975458 | A | G | 0.057 | 0.0214 | 0.0031 | 1.6E-09 | 48.9 |
| European | Targeting LDL receptors | *LDLR* | rs34243815 | T | C | 0.063 | -0.0232 | 0.0029 | 8.9E-12 | 62.5 |
| European | Targeting LDL receptors | *LDLR* | rs9967639 | T | A | 0.074 | 0.0222 | 0.0027 | 9.6E-13 | 68.4 |
| European | Targeting LDL receptors | *LDLR* | rs66466742 | T | C | 0.039 | -0.0514 | 0.0035 | 6.7E-36 | 210.2 |
| European | Targeting LDL receptors | *LDLR* | rs1433091 | A | G | 0.088 | 0.0305 | 0.0026 | 4.3E-24 | 137.7 |
| European | Mipomersen | *APOB* | rs62120800 | A | G | 0.013 | -0.1088 | 0.0062 | 5.7E-51 | 302.9 |
| European | Mipomersen | *APOB* | rs137978266 | G | A | 0.013 | 0.0818 | 0.0062 | 2.5E-30 | 176.0 |
| European | Mipomersen | *APOB* | rs79106033 | T | C | 0.026 | -0.0300 | 0.0044 | 5.8E-09 | 45.5 |
| European | Mipomersen | *APOB* | rs576306544 | A | C | 0.012 | -0.0578 | 0.0074 | 1.2E-11 | 61.8 |
| European | Mipomersen | *APOB* | rs1317821 | T | C | 0.253 | 0.0515 | 0.0016 | 1.1E-169 | 1035.5 |
| European | Mipomersen | *APOB* | rs76384951 | C | A | 0.076 | -0.0182 | 0.0027 | 9.8E-09 | 44.2 |
| European | Mipomersen | *APOB* | rs62122481 | A | C | 0.359 | 0.0705 | 0.0015 | <4.9E-324 | 2347.9 |
| European | Mipomersen | *APOB* | rs1042023 | C | G | 0.012 | 0.1359 | 0.0068 | 1.3E-67 | 405.5 |
| European | Mipomersen | *APOB* | rs533617 | C | T | 0.041 | -0.1261 | 0.0035 | 4.9E-215 | 1315.8 |

| | | | | | | | | | | |
|---|---|---|---|---|---|---|---|---|---|---|
| European | Mipomersen | *APOB* | rs497166 | T | C | 0.020 | -0.0959 | 0.0049 | 2.7E-63 | 378.9 |
| European | Mipomersen | *APOB* | rs184507838 | T | C | 0.010 | -0.1258 | 0.0069 | 2.8E-56 | 335.6 |
| European | Mipomersen | *APOB* | rs531819 | T | G | 0.147 | -0.1160 | 0.0020 | <4.9E-324 | 3441.1 |
| European | Mipomersen | *APOB* | rs72653053 | C | T | 0.011 | -0.0799 | 0.0068 | 3.5E-24 | 138.2 |
| European | Mipomersen | *APOB* | rs9282606 | T | G | 0.045 | 0.0649 | 0.0034 | 2.9E-59 | 354.0 |
| European | Mipomersen | *APOB* | rs3056574 | A | T | 0.137 | 0.0302 | 0.0026 | 3.7E-23 | 132.0 |
| European | Mipomersen | *APOB* | rs72902579 | C | T | 0.038 | -0.0975 | 0.0037 | 6.6E-115 | 697.3 |
| European | Mipomersen | *APOB* | rs200362396 | A | T | 0.344 | 0.0533 | 0.0032 | 3.8E-46 | 273.2 |
| European | Mipomersen | *APOB* | rs72780178 | C | A | 0.027 | 0.0608 | 0.0043 | 1.2E-33 | 196.4 |
| European | Mipomersen | *APOB* | rs78158108 | C | G | 0.074 | 0.0817 | 0.0027 | 5.9E-146 | 889.0 |
| European | Mipomersen | *APOB* | rs377122620 | G | T | 0.097 | 0.1046 | 0.0041 | 2.2E-105 | 638.5 |
| European | Mipomersen | *APOB* | rs145713446 | A | T | 0.006 | 0.0971 | 0.0096 | 2.6E-18 | 102.3 |
| European | Mipomersen | *APOB* | rs114185526 | T | C | 0.008 | -0.1056 | 0.0082 | 7.6E-29 | 166.8 |
| European | Mipomersen | *APOB* | rs17395484 | T | C | 0.022 | 0.0669 | 0.0049 | 4.7E-32 | 186.6 |
| European | Mipomersen | *APOB* | rs548774896 | G | T | 0.118 | 0.0667 | 0.0042 | 4.7E-42 | 248.0 |
| European | Mipomersen | *APOB* | rs72782175 | C | T | 0.014 | -0.0955 | 0.0060 | 2.4E-42 | 249.8 |
| European | Targeting ABCG5/8 | *ABCG5/8* | rs145288624 | T | C | 0.066 | 0.0281 | 0.0030 | 7.0E-16 | 87.5 |
| European | Targeting ABCG5/8 | *ABCG5/8* | rs78354019 | G | A | 0.056 | 0.0230 | 0.0031 | 8.9E-11 | 56.5 |
| European | Targeting ABCG5/8 | *ABCG5/8* | rs143706998 | C | A | 0.008 | -0.1129 | 0.0081 | 2.0E-33 | 194.9 |
| European | Targeting ABCG5/8 | *ABCG5/8* | rs576115488 | G | T | 0.012 | -0.0877 | 0.0082 | 1.6E-20 | 115.7 |
| European | Targeting ABCG5/8 | *ABCG5/8* | rs113140405 | T | C | 0.018 | -0.0504 | 0.0057 | 1.7E-14 | 79.0 |
| European | Targeting ABCG5/8 | *ABCG5/8* | rs10186086 | A | T | 0.194 | -0.0216 | 0.0018 | 5.8E-26 | 149.1 |
| European | Targeting ABCG5/8 | *ABCG5/8* | rs531043156 | G | T | 0.018 | -0.0907 | 0.0076 | 3.9E-25 | 144.1 |

| | | | | | | | | | | |
|---|---|---|---|---|---|---|---|---|---|---|
| European | Targeting ABCG5/8 | *ABCG5/8* | rs139029940 | A | C | 0.152 | 0.0348 | 0.0020 | 1.6E-51 | 306.2 |
| European | Targeting ABCG5/8 | *ABCG5/8* | rs4131228 | G | A | 0.021 | 0.0415 | 0.0052 | 4.5E-12 | 64.3 |
| European | Targeting ABCG5/8 | *ABCG5/8* | rs182201537 | G | A | 0.009 | 0.0835 | 0.0078 | 2.7E-20 | 114.5 |
| European | Targeting ABCG5/8 | *ABCG5/8* | rs116487096 | A | T | 0.039 | -0.0288 | 0.0037 | 1.8E-11 | 60.6 |
| European | Targeting ABCG5/8 | *ABCG5/8* | rs75331444 | A | G | 0.066 | -0.1146 | 0.0028 | 3.6E-278 | 1706.0 |
| European | Targeting ABCG5/8 | *ABCG5/8* | rs55726838 | A | G | 0.010 | 0.0784 | 0.0075 | 2.3E-19 | 108.8 |
| European | Targeting ABCG5/8 | *ABCG5/8* | rs4299376 | G | T | 0.309 | 0.0700 | 0.0015 | <4.9E-324 | 2155.6 |
| European | Targeting ABCG5/8 | *ABCG5/8* | rs67734975 | G | C | 0.045 | 0.0525 | 0.0033 | 1.0E-42 | 252.1 |
| European | Targeting ABCG5/8 | *ABCG5/8* | rs10221914 | T | C | 0.028 | 0.0296 | 0.0045 | 1.2E-08 | 43.6 |
| European | Targeting ABCG5/8 | *ABCG5/8* | rs4148216 | T | C | 0.169 | -0.0281 | 0.0019 | 7.6E-39 | 228.3 |
| European | Targeting ABCG5/8 | *ABCG5/8* | rs187356514 | C | T | 0.013 | 0.0507 | 0.0063 | 5.0E-12 | 64.0 |
| European | Targeting ABCG5/8 | *ABCG5/8* | rs13396356 | T | A | 0.058 | -0.0286 | 0.0030 | 2.2E-16 | 90.5 |
| European | Targeting ABCG5/8 | *ABCG5/8* | rs149109487 | C | G | 0.020 | -0.0414 | 0.0051 | 1.7E-12 | 66.9 |
| European | Targeting ABCG5/8 | *ABCG5/8* | rs75841075 | A | G | 0.023 | 0.0310 | 0.0046 | 7.6E-09 | 44.8 |
| European | Targeting ABCG5/8 | *ABCG5/8* | rs7573769 | G | A | 0.176 | -0.0181 | 0.0018 | 1.2E-17 | 98.2 |
| European | Targeting ABCG5/8 | *ABCG5/8* | rs111859423 | C | T | 0.032 | 0.0333 | 0.0043 | 1.4E-11 | 61.3 |
| European | Targeting ABCG5/8 | *ABCG5/8* | rs188238029 | T | A | 0.009 | 0.0514 | 0.0076 | 6.4E-09 | 45.3 |
| European | Targeting ABCG5/8 | *ABCG5/8* | rs62135111 | A | G | 0.143 | 0.0222 | 0.0020 | 3.0E-22 | 126.4 |
| East Asian | Statins | *HMGCR* | rs7717415 | G | A | 0.470 | 0.0408 | 0.0052 | 4.4E-14 | 62.1 |
| East Asian | Statins | *HMGCR* | rs78908099 | C | T | 0.042 | 0.0851 | 0.0128 | 2.2E-10 | 43.9 |
| East Asian | Statins | *HMGCR* | rs77679151 | T | C | 0.053 | -0.0698 | 0.0113 | 3.7E-09 | 37.9 |
| East Asian | Statins | *HMGCR* | rs6453131 | T | G | 0.498 | -0.0815 | 0.0050 | 4.3E-55 | 266.3 |
| East Asian | Statins | *HMGCR* | rs139488631 | C | T | 0.013 | -0.1584 | 0.0253 | 1.9E-09 | 39.3 |

| | | | | | | | | | | |
|---|---|---|---|---|---|---|---|---|---|---|
| East Asian | PCSK9 inhibitors | *PCSK9* | rs34232196 | T | C | 0.127 | -0.0607 | 0.0077 | 4.3E-14 | 62.1 |
| East Asian | PCSK9 inhibitors | *PCSK9* | rs28848281 | A | G | 0.425 | -0.0354 | 0.0058 | 5.7E-09 | 37.0 |
| East Asian | PCSK9 inhibitors | *PCSK9* | rs2495491 | T | G | 0.273 | -0.0371 | 0.0061 | 5.9E-09 | 36.9 |
| East Asian | PCSK9 inhibitors | *PCSK9* | rs151193009 | T | C | 0.009 | -0.4860 | 0.0333 | 2.3E-44 | 212.7 |
| East Asian | PCSK9 inhibitors | *PCSK9* | rs10888897 | T | C | 0.187 | -0.0604 | 0.0066 | 1.3E-18 | 84.6 |
| East Asian | PCSK9 inhibitors | *PCSK9* | rs17111652 | T | C | 0.060 | 0.0763 | 0.0107 | 1.0E-11 | 50.4 |
| East Asian | PCSK9 inhibitors | *PCSK9* | rs6663252 | C | T | 0.120 | -0.0751 | 0.0078 | 3.5E-20 | 92.2 |
| East Asian | Targeting LDL receptors | *LDLR* | rs145960625 | A | G | 0.008 | -0.2053 | 0.0308 | 1.7E-10 | 44.4 |
| East Asian | Targeting LDL receptors | *LDLR* | rs67475684 | T | A | 0.162 | 0.0713 | 0.0069 | 4.5E-23 | 106.6 |
| East Asian | Targeting LDL receptors | *LDLR* | rs5927 | A | G | 0.057 | -0.0859 | 0.0116 | 1.5E-12 | 54.5 |
| East Asian | Targeting LDL receptors | *LDLR* | rs117373926 | G | T | 0.112 | 0.0514 | 0.0085 | 6.5E-09 | 36.7 |
| East Asian | Targeting LDL receptors | *LDLR* | rs2738464 | G | C | 0.283 | -0.1080 | 0.0059 | 8.3E-68 | 329.9 |
| East Asian | Targeting LDL receptors | *LDLR* | rs117523171 | G | T | 0.327 | 0.0398 | 0.0067 | 1.3E-08 | 35.2 |
| East Asian | Targeting LDL receptors | *LDLR* | rs183241760 | A | G | 0.018 | -0.1617 | 0.0200 | 9.2E-15 | 65.4 |
| East Asian | Targeting LDL receptors | *LDLR* | rs442363 | A | G | 0.408 | 0.0371 | 0.0056 | 2.2E-10 | 43.9 |
| East Asian | Targeting LDL receptors | *LDLR* | rs17699089 | G | A | 0.261 | -0.0620 | 0.0057 | 3.0E-25 | 117.4 |
| East Asian | Mipomersen | *APOB* | rs76684042 | T | A | 0.024 | 0.1263 | 0.0204 | 2.8E-09 | 38.4 |
| East Asian | Mipomersen | *APOB* | rs76693756 | T | C | 0.017 | 0.1183 | 0.0196 | 6.8E-09 | 36.6 |
| East Asian | Mipomersen | *APOB* | rs13306194 | A | G | 0.114 | -0.1235 | 0.0079 | 8.6E-51 | 244.8 |
| East Asian | Mipomersen | *APOB* | rs589566 | A | G | 0.216 | 0.0649 | 0.0063 | 6.0E-23 | 106.0 |
| East Asian | Mipomersen | *APOB* | rs528148801 | A | C | 0.009 | -0.1590 | 0.0259 | 3.8E-09 | 37.8 |

ABCG5/8, adenosine triphosphate (ATP)-binding cassette transporters G5/8; LDL, low-density lipoprotein; PCSK9, proprotein convertase subtilisin/kexin type 9.

Supplemental Table 4. SNP-specific estimates for genetic predictors for plasma LDL-cholesterol on plasma LDL-cholesterol (standard deviation) in people of European and East Asian ancestry.

| Ancestry | SNP | Effect allele | Other allele | Effect allele frequency | Beta | SE | *P* value | F-statistic |
|---|---|---|---|---|---|---|---|---|
| European | rs1123571 | A | G | 0.469 | -0.0110 | 0.0014 | 3.0E-11 | 59.3 |
| European | rs1497406 | A | G | 0.416 | -0.0136 | 0.0014 | 7.5E-17 | 93.4 |
| European | rs10903129 | A | G | 0.452 | -0.0258 | 0.0014 | 5.2E-58 | 346.3 |
| European | rs12094989 | T | C | 0.215 | -0.0129 | 0.0017 | 5.2E-11 | 57.9 |
| European | rs114165349 | C | G | 0.023 | 0.0835 | 0.0046 | 8.4E-55 | 326.5 |
| European | rs8681 | A | G | 0.256 | -0.0112 | 0.0016 | 1.4E-09 | 49.3 |
| European | rs213499 | C | A | 0.468 | -0.0102 | 0.0014 | 3.1E-10 | 53.2 |
| European | rs374459115 | A | G | 0.017 | 0.0683 | 0.0068 | 2.8E-18 | 102.2 |
| European | rs11591147 | T | G | 0.016 | -0.4337 | 0.0054 | <4.9E-324 | 6388.7 |
| European | rs693668 | G | A | 0.359 | -0.0565 | 0.0015 | 2.6E-243 | 1490.6 |
| European | rs11206517 | G | T | 0.037 | 0.0889 | 0.0037 | 7.6E-95 | 573.4 |
| European | rs6588577 | A | C | 0.066 | -0.0219 | 0.0028 | 1.9E-11 | 60.5 |
| European | rs7534572 | C | G | 0.329 | -0.0429 | 0.0015 | 2.2E-127 | 774.4 |
| European | rs2391159 | T | C | 0.207 | -0.0234 | 0.0017 | 6.6E-32 | 185.6 |
| European | rs35271870 | C | T | 0.084 | -0.1171 | 0.0025 | <4.9E-324 | 2164.6 |
| European | rs141521383 | C | G | 0.023 | 0.0374 | 0.0050 | 1.1E-10 | 55.9 |
| European | rs35358959 | A | G | 0.081 | -0.1051 | 0.0026 | 7.4E-276 | 1691.7 |
| European | rs17036094 | C | A | 0.012 | -0.1143 | 0.0064 | 2.4E-53 | 317.5 |
| European | rs61798348 | T | A | 0.019 | -0.0345 | 0.0052 | 9.6E-09 | 44.2 |
| European | rs267733 | G | A | 0.156 | -0.0187 | 0.0019 | 2.2E-17 | 96.7 |
| European | rs4390169 | A | G | 0.485 | 0.0120 | 0.0014 | 8.8E-14 | 74.7 |
| European | rs115383270 | A | G | 0.073 | 0.0193 | 0.0027 | 1.3E-09 | 49.4 |
| European | rs6682862 | A | G | 0.164 | -0.0137 | 0.0019 | 3.4E-10 | 52.9 |
| European | rs1689801 | A | G | 0.325 | 0.0144 | 0.0015 | 3.9E-17 | 95.2 |
| European | rs2296288 | T | C | 0.437 | -0.0113 | 0.0014 | 3.8E-12 | 64.8 |
| European | rs1434282 | C | T | 0.277 | -0.0109 | 0.0016 | 2.1E-09 | 48.2 |
| European | rs2642438 | A | G | 0.294 | -0.0269 | 0.0015 | 3.2E-52 | 310.6 |
| European | rs12138866 | G | A | 0.040 | -0.0240 | 0.0037 | 1.5E-08 | 43.1 |
| European | rs7544869 | A | T | 0.453 | -0.0122 | 0.0014 | 7.4E-14 | 75.2 |
| European | rs553427 | C | T | 0.473 | -0.0403 | 0.0014 | 7.0E-135 | 820.6 |
| European | rs10910522 | A | G | 0.429 | -0.0119 | 0.0014 | 4.6E-13 | 70.3 |
| European | rs7512010 | A | T | 0.193 | -0.0183 | 0.0018 | 1.6E-18 | 103.5 |

| | | | | | | | | |
|---|---|---|---|---|---|---|---|---|
| European | rs3935011 | C | T | 0.480 | 0.0089 | 0.0014 | 3.6E-08 | 40.7 |
| European | rs3820897 | T | C | 0.192 | 0.0159 | 0.0018 | 8.2E-15 | 81.0 |
| European | rs67269656 | T | C | 0.265 | -0.0104 | 0.0016 | 1.3E-08 | 43.5 |
| European | rs4465730 | A | G | 0.219 | 0.0146 | 0.0017 | 6.2E-14 | 75.7 |
| European | rs77370158 | G | A | 0.077 | 0.0298 | 0.0027 | 9.4E-22 | 123.4 |
| European | rs115692156 | G | A | 0.010 | -0.0749 | 0.0074 | 1.8E-18 | 103.3 |
| European | rs62122481 | A | C | 0.359 | 0.0705 | 0.0015 | <4.9E-324 | 2347.9 |
| European | rs12720796 | C | A | 0.021 | 0.0692 | 0.0049 | 6.8E-34 | 197.8 |
| European | rs72902579 | C | T | 0.038 | -0.0975 | 0.0037 | 6.6E-115 | 697.3 |
| European | rs11891554 | A | G | 0.053 | -0.0234 | 0.0032 | 2.0E-10 | 54.4 |
| European | rs2374569 | T | C | 0.369 | -0.0092 | 0.0014 | 3.8E-08 | 40.6 |
| European | rs185263492 | A | T | 0.159 | 0.0306 | 0.0020 | 1.6E-38 | 226.2 |
| European | rs4299376 | G | T | 0.309 | 0.0700 | 0.0015 | <4.9E-324 | 2155.6 |
| European | rs6709904 | G | A | 0.109 | -0.0472 | 0.0022 | 3.8E-76 | 458.0 |
| European | rs6741740 | A | G | 0.241 | 0.0130 | 0.0016 | 5.2E-12 | 64.0 |
| European | rs4671050 | T | G | 0.320 | -0.0224 | 0.0015 | 8.3E-38 | 221.9 |
| European | rs954680 | C | G | 0.298 | -0.0124 | 0.0015 | 4.2E-12 | 64.5 |
| European | rs11887443 | T | G | 0.263 | -0.0125 | 0.0018 | 9.5E-10 | 50.3 |
| European | rs2970902 | C | G | 0.342 | 0.0101 | 0.0015 | 3.1E-09 | 47.1 |
| European | rs113313551 | G | C | 0.148 | -0.0127 | 0.0020 | 2.0E-08 | 42.3 |
| European | rs1992172 | G | A | 0.194 | 0.0195 | 0.0018 | 1.1E-21 | 122.9 |
| European | rs150474434 | A | G | 0.095 | -0.0423 | 0.0024 | 4.9E-53 | 315.6 |
| European | rs17050272 | A | G | 0.421 | -0.0228 | 0.0014 | 3.2E-42 | 249.0 |
| European | rs1375131 | C | T | 0.313 | 0.0196 | 0.0017 | 1.3E-22 | 128.7 |
| European | rs12614487 | T | C | 0.074 | -0.0266 | 0.0026 | 3.0E-18 | 102.0 |
| European | rs10184004 | T | C | 0.413 | -0.0105 | 0.0014 | 1.4E-10 | 55.3 |
| European | rs10184673 | G | A | 0.405 | 0.0220 | 0.0014 | 1.4E-40 | 239.0 |
| European | rs12693968 | A | G | 0.258 | 0.0213 | 0.0016 | 1.6E-30 | 177.1 |
| European | rs3814365 | T | C | 0.443 | -0.0137 | 0.0014 | 5.7E-17 | 94.1 |
| European | rs1250259 | T | A | 0.264 | -0.0177 | 0.0016 | 1.2E-21 | 122.6 |
| European | rs78058190 | A | G | 0.052 | 0.0240 | 0.0036 | 1.0E-08 | 44.1 |
| European | rs6431630 | A | G | 0.105 | 0.0241 | 0.0023 | 2.9E-20 | 114.2 |
| European | rs146194062 | T | C | 0.012 | -0.0702 | 0.0070 | 8.1E-18 | 99.3 |
| European | rs13076933 | G | T | 0.257 | -0.0245 | 0.0016 | 6.3E-40 | 234.9 |
| European | rs6792725 | A | G | 0.319 | 0.0153 | 0.0016 | 2.9E-17 | 95.9 |
| European | rs9837622 | A | T | 0.073 | -0.0371 | 0.0027 | 2.8E-32 | 187.9 |

| | | | | | | | | |
|---|---|---|---|---|---|---|---|---|
| European | rs71311871 | G | A | 0.081 | -0.0380 | 0.0025 | 1.3E-38 | 226.8 |
| European | rs55921103 | G | T | 0.359 | -0.0126 | 0.0015 | 1.8E-13 | 72.8 |
| European | rs9837149 | C | G | 0.153 | 0.0189 | 0.0019 | 2.2E-17 | 96.6 |
| European | rs11719535 | C | T | 0.244 | -0.0118 | 0.0016 | 4.4E-10 | 52.3 |
| European | rs55732372 | A | G | 0.076 | 0.0174 | 0.0027 | 2.0E-08 | 42.3 |
| European | rs74341202 | A | G | 0.050 | -0.0426 | 0.0032 | 5.2E-30 | 174.0 |
| European | rs523118 | T | G | 0.211 | -0.0144 | 0.0018 | 2.2E-12 | 66.3 |
| European | rs9653945 | A | G | 0.347 | -0.0130 | 0.0015 | 1.4E-14 | 79.5 |
| European | rs6441313 | A | G | 0.463 | 0.0126 | 0.0014 | 2.0E-14 | 78.6 |
| European | rs56118251 | G | A | 0.157 | 0.0137 | 0.0019 | 4.1E-10 | 52.5 |
| European | rs13108218 | A | G | 0.385 | 0.0183 | 0.0015 | 5.2E-27 | 155.6 |
| European | rs4234798 | T | G | 0.390 | -0.0110 | 0.0014 | 2.0E-11 | 60.3 |
| European | rs79623641 | A | G | 0.068 | -0.0179 | 0.0028 | 4.4E-08 | 40.3 |
| European | rs112575086 | T | C | 0.120 | -0.0139 | 0.0021 | 1.8E-08 | 42.7 |
| European | rs146674238 | T | A | 0.014 | 0.0472 | 0.0065 | 4.9E-10 | 52.0 |
| European | rs34707604 | C | T | 0.248 | 0.0318 | 0.0019 | 1.1E-48 | 288.9 |
| European | rs149314105 | T | C | 0.028 | -0.0339 | 0.0050 | 5.2E-09 | 45.8 |
| European | rs72663045 | G | T | 0.020 | 0.0403 | 0.0050 | 5.1E-12 | 64.0 |
| European | rs342467 | T | C | 0.399 | -0.0102 | 0.0014 | 6.6E-10 | 51.2 |
| European | rs11499828 | C | T | 0.108 | 0.0165 | 0.0023 | 2.7E-10 | 53.6 |
| European | rs28497720 | T | C | 0.251 | -0.0175 | 0.0016 | 2.5E-21 | 120.8 |
| European | rs17617028 | A | G | 0.217 | 0.0133 | 0.0017 | 8.7E-12 | 62.6 |
| European | rs138204164 | G | C | 0.132 | -0.0145 | 0.0021 | 1.3E-09 | 49.5 |
| European | rs41280463 | A | G | 0.167 | -0.0152 | 0.0019 | 1.6E-12 | 67.1 |
| European | rs116692022 | G | A | 0.013 | 0.0455 | 0.0065 | 1.2E-09 | 49.7 |
| European | rs72701754 | T | A | 0.404 | 0.0092 | 0.0014 | 3.6E-08 | 40.7 |
| European | rs116734477 | T | C | 0.038 | -0.0511 | 0.0037 | 1.9E-33 | 195.1 |
| European | rs3010275 | G | T | 0.210 | -0.0183 | 0.0017 | 1.6E-20 | 115.9 |
| European | rs1423527 | A | C | 0.399 | 0.0653 | 0.0014 | <4.9E-324 | 2127.3 |
| European | rs6869845 | C | T | 0.451 | -0.0167 | 0.0014 | 5.7E-25 | 143.1 |
| European | rs2706381 | T | C | 0.196 | -0.0195 | 0.0018 | 7.0E-22 | 124.1 |
| European | rs13153174 | G | C | 0.214 | -0.0108 | 0.0017 | 3.8E-08 | 40.6 |
| European | rs11167778 | T | C | 0.107 | 0.0218 | 0.0023 | 6.2E-17 | 93.9 |
| European | rs543486395 | A | C | 0.023 | -0.0488 | 0.0060 | 2.4E-12 | 66.0 |
| European | rs12657266 | C | T | 0.365 | -0.0353 | 0.0014 | 9.0E-100 | 603.8 |
| European | rs13360569 | T | C | 0.430 | -0.0098 | 0.0014 | 2.1E-09 | 48.2 |

| | | | | | | | | |
|---|---|---|---|---|---|---|---|---|
| European | rs352942 | A | G | 0.252 | -0.0104 | 0.0016 | 3.8E-08 | 40.6 |
| European | rs147539187 | G | C | 0.069 | -0.0231 | 0.0028 | 6.0E-13 | 69.6 |
| European | rs2235215 | C | T | 0.321 | -0.0318 | 0.0015 | 4.3E-76 | 457.7 |
| European | rs1800562 | A | G | 0.068 | -0.0572 | 0.0028 | 2.2E-71 | 428.7 |
| European | rs1061537 | A | G | 0.444 | 0.0136 | 0.0016 | 2.1E-13 | 72.4 |
| European | rs28732146 | A | T | 0.196 | 0.0140 | 0.0018 | 2.0E-11 | 60.4 |
| European | rs6689 | G | A | 0.203 | 0.0390 | 0.0019 | 2.0E-70 | 422.8 |
| European | rs114863007 | A | G | 0.097 | -0.0267 | 0.0024 | 1.1E-22 | 129.0 |
| European | rs62406547 | C | A | 0.496 | -0.0128 | 0.0016 | 1.1E-12 | 68.0 |
| European | rs17665178 | G | C | 0.302 | -0.0159 | 0.0016 | 1.6E-18 | 103.6 |
| European | rs12662589 | C | G | 0.259 | 0.0153 | 0.0016 | 7.8E-17 | 93.3 |
| European | rs9496567 | A | G | 0.236 | -0.0210 | 0.0016 | 3.4E-28 | 162.8 |
| European | rs62419249 | A | G | 0.478 | 0.0121 | 0.0014 | 1.0E-13 | 74.3 |
| European | rs4946713 | A | C | 0.447 | -0.0112 | 0.0014 | 4.9E-12 | 64.1 |
| European | rs1556857 | C | T | 0.407 | -0.0169 | 0.0014 | 4.4E-25 | 143.7 |
| European | rs72971192 | C | T | 0.235 | -0.0118 | 0.0017 | 1.4E-09 | 49.3 |
| European | rs141783576 | C | G | 0.069 | 0.0240 | 0.0031 | 1.5E-11 | 61.2 |
| European | rs9399137 | C | T | 0.262 | -0.0256 | 0.0016 | 4.4E-44 | 260.5 |
| European | rs76933614 | C | T | 0.077 | 0.0232 | 0.0027 | 7.4E-14 | 75.2 |
| European | rs112170089 | A | G | 0.018 | 0.0521 | 0.0059 | 3.6E-14 | 77.1 |
| European | rs12208357 | T | C | 0.070 | 0.0645 | 0.0027 | 1.9E-94 | 571.0 |
| European | rs146534110 | T | G | 0.012 | 0.0761 | 0.0066 | 4.9E-23 | 131.2 |
| European | rs144833821 | C | T | 0.009 | -0.0498 | 0.0076 | 1.6E-08 | 42.9 |
| European | rs117733303 | G | A | 0.018 | 0.1453 | 0.0053 | 1.2E-123 | 751.2 |
| European | rs10455872 | G | A | 0.069 | 0.1138 | 0.0027 | 3.2E-281 | 1724.9 |
| European | rs12055389 | T | C | 0.057 | -0.0278 | 0.0030 | 1.6E-15 | 85.3 |
| European | rs10272002 | G | A | 0.210 | -0.0223 | 0.0017 | 2.9E-29 | 169.4 |
| European | rs34927723 | T | C | 0.157 | -0.0134 | 0.0019 | 2.3E-09 | 48.0 |
| European | rs55696093 | G | A | 0.211 | 0.0382 | 0.0017 | 3.1E-83 | 501.8 |
| European | rs896311 | G | A | 0.300 | -0.0166 | 0.0015 | 7.4E-21 | 117.9 |
| European | rs56001710 | A | T | 0.423 | -0.0137 | 0.0017 | 7.1E-12 | 63.1 |
| European | rs12533280 | T | C | 0.195 | 0.0180 | 0.0018 | 9.4E-19 | 105.0 |
| European | rs7808613 | G | C | 0.245 | 0.0113 | 0.0016 | 2.7E-09 | 47.6 |
| European | rs17725246 | C | T | 0.195 | 0.0428 | 0.0018 | 1.3E-94 | 572.0 |
| European | rs799157 | T | C | 0.036 | 0.0318 | 0.0039 | 1.4E-12 | 67.3 |
| European | rs1057868 | T | C | 0.288 | 0.0115 | 0.0015 | 7.7E-11 | 56.9 |

| | | | | | | | | |
|---|---|---|---|---|---|---|---|---|
| European | rs1014283 | A | C | 0.183 | -0.0136 | 0.0018 | 6.1E-11 | 57.5 |
| European | rs6967728 | A | G | 0.183 | -0.0179 | 0.0018 | 3.5E-17 | 95.4 |
| European | rs564449 | T | G | 0.115 | 0.0267 | 0.0022 | 3.9E-26 | 150.2 |
| European | rs1838931 | T | C | 0.312 | 0.0113 | 0.0015 | 1.5E-10 | 55.2 |
| European | rs4374942 | C | T | 0.081 | 0.0200 | 0.0026 | 1.6E-11 | 61.1 |
| European | rs2936512 | T | C | 0.326 | -0.0119 | 0.0015 | 4.0E-12 | 64.6 |
| European | rs9987289 | A | G | 0.092 | -0.0599 | 0.0024 | 1.6E-103 | 627.1 |
| European | rs10108282 | A | T | 0.212 | 0.0139 | 0.0017 | 2.3E-12 | 66.1 |
| European | rs13255048 | A | G | 0.141 | 0.0210 | 0.0020 | 9.6E-20 | 111.1 |
| European | rs151150389 | C | T | 0.034 | 0.0298 | 0.0038 | 1.9E-11 | 60.5 |
| European | rs117139027 | A | G | 0.013 | -0.0783 | 0.0064 | 7.3E-26 | 148.5 |
| European | rs28615248 | C | T | 0.196 | 0.0244 | 0.0018 | 4.0E-33 | 193.1 |
| European | rs9297994 | G | A | 0.338 | 0.0355 | 0.0015 | 4.0E-97 | 587.5 |
| European | rs12114596 | T | C | 0.347 | 0.0117 | 0.0016 | 3.4E-10 | 53.0 |
| European | rs62509311 | T | A | 0.279 | -0.0153 | 0.0016 | 3.4E-17 | 95.5 |
| European | rs2737245 | T | G | 0.277 | -0.0254 | 0.0016 | 3.2E-45 | 267.5 |
| European | rs13249867 | T | G | 0.136 | -0.0207 | 0.0027 | 2.7E-11 | 59.6 |
| European | rs28601761 | G | C | 0.422 | -0.0649 | 0.0014 | <4.9E-324 | 2092.2 |
| European | rs11787335 | T | C | 0.355 | 0.0231 | 0.0015 | 3.8E-42 | 248.6 |
| European | rs3780181 | G | A | 0.070 | -0.0359 | 0.0028 | 3.8E-29 | 168.7 |
| European | rs28498684 | A | G | 0.400 | 0.0132 | 0.0014 | 1.3E-15 | 85.8 |
| European | rs12551960 | T | C | 0.079 | 0.0350 | 0.0026 | 4.3E-30 | 174.5 |
| European | rs7864568 | A | G | 0.316 | -0.0156 | 0.0016 | 7.4E-18 | 99.6 |
| European | rs1571791 | T | C | 0.380 | 0.0139 | 0.0014 | 8.1E-17 | 93.2 |
| European | rs9410207 | C | T | 0.066 | -0.0200 | 0.0028 | 9.8E-10 | 50.2 |
| European | rs2066714 | C | T | 0.124 | 0.0190 | 0.0021 | 5.2E-15 | 82.2 |
| European | rs11789603 | T | C | 0.101 | 0.0236 | 0.0023 | 2.2E-18 | 102.7 |
| European | rs2740488 | C | A | 0.261 | -0.0249 | 0.0016 | 4.6E-42 | 248.1 |
| European | rs56294298 | A | G | 0.085 | -0.0183 | 0.0026 | 1.4E-09 | 49.2 |
| European | rs6478851 | A | G | 0.236 | -0.0132 | 0.0016 | 4.1E-12 | 64.5 |
| European | rs2519093 | T | C | 0.188 | 0.0724 | 0.0018 | 5.8E-278 | 1704.7 |
| European | rs76643124 | A | G | 0.024 | -0.0289 | 0.0045 | 3.0E-08 | 41.3 |
| European | rs13301660 | T | C | 0.270 | -0.0156 | 0.0016 | 8.2E-18 | 99.3 |
| European | rs7903259 | G | C | 0.416 | 0.0163 | 0.0014 | 4.3E-23 | 131.6 |
| European | rs1896995 | C | T | 0.498 | -0.0122 | 0.0014 | 8.7E-14 | 74.7 |
| European | rs17476364 | C | T | 0.099 | -0.0268 | 0.0023 | 3.0E-23 | 132.5 |

| | | | | | | | | |
|---|---|---|---|---|---|---|---|---|
| European | rs1870140 | A | G | 0.157 | -0.0125 | 0.0019 | 2.2E-08 | 42.0 |
| European | rs2068888 | A | G | 0.451 | -0.0180 | 0.0014 | 7.3E-29 | 166.9 |
| European | rs61886346 | T | C | 0.061 | -0.0192 | 0.0029 | 1.2E-08 | 43.6 |
| European | rs603424 | A | G | 0.169 | 0.0159 | 0.0019 | 3.7E-13 | 70.9 |
| European | rs2792751 | T | C | 0.281 | 0.0239 | 0.0015 | 2.7E-41 | 243.3 |
| European | rs60847460 | T | C | 0.140 | -0.0182 | 0.0020 | 3.5E-15 | 83.2 |
| European | rs2301179 | A | G | 0.492 | -0.0159 | 0.0014 | 8.2E-23 | 129.8 |
| European | rs7904973 | G | T | 0.424 | -0.0210 | 0.0014 | 7.0E-38 | 222.4 |
| European | rs7124487 | T | C | 0.186 | -0.0116 | 0.0018 | 4.1E-08 | 40.4 |
| European | rs7108486 | C | T | 0.024 | -0.0322 | 0.0046 | 1.3E-09 | 49.4 |
| European | rs12271333 | C | A | 0.134 | 0.0165 | 0.0020 | 2.7E-12 | 65.7 |
| European | rs11601507 | A | C | 0.070 | 0.0404 | 0.0032 | 4.8E-28 | 161.9 |
| European | rs10832956 | T | C | 0.272 | -0.0208 | 0.0016 | 1.0E-30 | 178.4 |
| European | rs61882680 | T | C | 0.030 | -0.0280 | 0.0041 | 4.3E-09 | 46.3 |
| European | rs174547 | C | T | 0.346 | -0.0429 | 0.0015 | 6.4E-143 | 870.3 |
| European | rs77631946 | A | C | 0.093 | -0.0187 | 0.0025 | 5.2E-11 | 57.9 |
| European | rs1638586 | A | G | 0.080 | -0.0196 | 0.0026 | 5.2E-11 | 57.9 |
| European | rs78643851 | T | G | 0.021 | 0.0348 | 0.0054 | 2.5E-08 | 41.7 |
| European | rs11226108 | C | G | 0.191 | -0.0143 | 0.0018 | 7.9E-12 | 62.8 |
| European | rs964184 | G | C | 0.135 | 0.0552 | 0.0020 | 3.5E-123 | 748.4 |
| European | rs141469619 | G | A | 0.009 | 0.0725 | 0.0080 | 3.8E-15 | 83.0 |
| European | rs12970 | A | G | 0.061 | -0.0268 | 0.0029 | 1.6E-15 | 85.4 |
| European | rs4307732 | A | G | 0.108 | 0.0480 | 0.0022 | 7.9E-78 | 468.5 |
| European | rs35882350 | G | A | 0.256 | 0.0182 | 0.0016 | 5.4E-22 | 124.8 |
| European | rs117233107 | A | G | 0.015 | -0.0459 | 0.0060 | 3.7E-11 | 58.8 |
| European | rs75667995 | C | T | 0.068 | -0.0291 | 0.0027 | 3.4E-20 | 113.9 |
| European | rs11175540 | A | T | 0.066 | 0.0261 | 0.0028 | 2.7E-15 | 83.9 |
| European | rs2250751 | A | G | 0.341 | -0.0165 | 0.0015 | 4.7E-22 | 125.2 |
| European | rs7300192 | A | G | 0.365 | 0.0135 | 0.0015 | 2.5E-15 | 84.1 |
| European | rs74090765 | G | T | 0.173 | 0.0145 | 0.0018 | 1.0E-11 | 62.2 |
| European | rs61754230 | T | C | 0.016 | 0.0491 | 0.0057 | 1.6E-13 | 73.2 |
| European | rs12306780 | T | A | 0.342 | 0.0116 | 0.0015 | 9.8E-12 | 62.3 |
| European | rs1515565 | A | G | 0.482 | -0.0093 | 0.0014 | 9.0E-09 | 44.4 |
| European | rs11837065 | T | C | 0.366 | -0.0111 | 0.0015 | 7.2E-11 | 57.0 |
| European | rs3184504 | T | C | 0.473 | -0.0234 | 0.0014 | 1.3E-47 | 282.2 |
| European | rs1169288 | C | A | 0.322 | 0.0359 | 0.0015 | 1.1E-95 | 578.7 |

| | | | | | | | | |
|---|---|---|---|---|---|---|---|---|
| European | rs2247139 | A | G | 0.132 | 0.0132 | 0.0020 | 2.4E-08 | 41.8 |
| European | rs2451322 | G | A | 0.405 | -0.0106 | 0.0014 | 1.1E-10 | 55.9 |
| European | rs11057841 | T | C | 0.141 | 0.0219 | 0.0020 | 9.0E-21 | 117.3 |
| European | rs75588192 | A | G | 0.140 | 0.0165 | 0.0021 | 2.2E-11 | 60.2 |
| European | rs7327867 | G | A | 0.474 | 0.0199 | 0.0014 | 1.4E-34 | 202.1 |
| European | rs208432 | C | T | 0.386 | 0.0107 | 0.0014 | 1.3E-10 | 55.4 |
| European | rs17532371 | G | C | 0.071 | -0.0194 | 0.0027 | 7.3E-10 | 50.9 |
| European | rs9592980 | G | A | 0.415 | -0.0095 | 0.0014 | 8.0E-09 | 44.7 |
| European | rs7330899 | A | G | 0.244 | 0.0143 | 0.0016 | 1.8E-14 | 79.0 |
| European | rs551473284 | T | C | 0.366 | -0.0152 | 0.0017 | 3.4E-15 | 83.3 |
| European | rs6602909 | C | T | 0.330 | 0.0212 | 0.0015 | 1.3E-33 | 196.1 |
| European | rs12016920 | C | T | 0.193 | -0.0172 | 0.0018 | 2.0E-16 | 90.8 |
| European | rs11621792 | T | C | 0.449 | 0.0220 | 0.0014 | 1.6E-40 | 238.6 |
| European | rs11846704 | T | C | 0.267 | -0.0138 | 0.0016 | 3.4E-14 | 77.2 |
| European | rs17101394 | A | G | 0.160 | 0.0154 | 0.0019 | 3.4E-12 | 65.1 |
| European | rs7157399 | T | C | 0.140 | -0.0254 | 0.0020 | 8.8E-28 | 160.3 |
| European | rs13379043 | C | T | 0.271 | -0.0165 | 0.0016 | 9.1E-20 | 111.2 |
| European | rs34752362 | A | G | 0.443 | -0.0096 | 0.0014 | 3.9E-09 | 46.6 |
| European | rs17776811 | A | C | 0.397 | 0.0094 | 0.0014 | 1.2E-08 | 43.6 |
| European | rs28929474 | T | C | 0.018 | 0.0528 | 0.0052 | 1.6E-18 | 103.6 |
| European | rs17580 | A | T | 0.039 | 0.0498 | 0.0036 | 1.3E-32 | 189.9 |
| European | rs28375625 | A | C | 0.461 | -0.0091 | 0.0014 | 2.4E-08 | 41.9 |
| European | rs79391862 | C | A | 0.020 | -0.0639 | 0.0053 | 3.8E-25 | 144.2 |
| European | rs148086620 | G | A | 0.042 | 0.0297 | 0.0035 | 4.6E-13 | 70.3 |
| European | rs686958 | T | A | 0.181 | 0.0115 | 0.0018 | 4.0E-08 | 40.5 |
| European | rs11636087 | C | T | 0.298 | 0.0144 | 0.0015 | 9.5E-16 | 86.7 |
| European | rs12917376 | C | T | 0.423 | -0.0126 | 0.0014 | 2.2E-14 | 78.3 |
| European | rs8029797 | A | T | 0.313 | -0.0095 | 0.0015 | 4.0E-08 | 40.5 |
| European | rs12445804 | A | G | 0.076 | 0.0326 | 0.0027 | 1.0E-25 | 147.7 |
| European | rs35468353 | G | A | 0.377 | 0.0119 | 0.0014 | 7.8E-13 | 68.9 |
| European | rs247617 | A | C | 0.321 | -0.0367 | 0.0015 | 3.5E-101 | 612.5 |
| European | rs4788817 | C | T | 0.378 | 0.0118 | 0.0014 | 2.3E-12 | 66.1 |
| European | rs34042070 | G | C | 0.191 | 0.0573 | 0.0018 | 8.5E-173 | 1054.8 |
| European | rs7202323 | G | T | 0.228 | -0.0229 | 0.0017 | 1.8E-32 | 189.1 |
| European | rs7404072 | C | T | 0.286 | -0.0103 | 0.0016 | 1.0E-08 | 44.0 |
| European | rs67890964 | C | T | 0.381 | -0.0183 | 0.0015 | 1.3E-26 | 153.2 |

| | | | | | | | | |
|---|---|---|---|---|---|---|---|---|
| European | rs539705186 | G | T | 0.083 | 0.0209 | 0.0030 | 3.0E-09 | 47.3 |
| European | rs55714927 | T | C | 0.189 | -0.0349 | 0.0018 | 8.3E-61 | 363.5 |
| European | rs150688657 | A | G | 0.103 | 0.0216 | 0.0023 | 9.3E-16 | 86.7 |
| European | rs28811342 | C | T | 0.197 | 0.0126 | 0.0017 | 5.5E-10 | 51.7 |
| European | rs704 | A | G | 0.477 | 0.0189 | 0.0014 | 1.0E-31 | 184.5 |
| European | rs117765227 | T | C | 0.029 | -0.0311 | 0.0042 | 1.0E-10 | 56.1 |
| European | rs62070652 | T | C | 0.266 | 0.0134 | 0.0016 | 2.0E-13 | 72.5 |
| European | rs525767 | A | G | 0.346 | 0.0098 | 0.0015 | 7.2E-09 | 45.0 |
| European | rs12943633 | T | C | 0.081 | -0.0178 | 0.0026 | 2.0E-09 | 48.4 |
| European | rs12949918 | C | T | 0.427 | 0.0089 | 0.0014 | 4.2E-08 | 40.4 |
| European | rs72836561 | T | C | 0.030 | -0.0298 | 0.0040 | 2.1E-10 | 54.2 |
| European | rs12603290 | T | C | 0.495 | 0.0268 | 0.0014 | 4.1E-58 | 346.9 |
| European | rs1292069 | C | T | 0.454 | -0.0098 | 0.0014 | 1.4E-09 | 49.2 |
| European | rs1801689 | C | A | 0.026 | 0.0908 | 0.0043 | 1.5E-74 | 448.3 |
| European | rs78186330 | A | G | 0.208 | 0.0163 | 0.0018 | 1.4E-15 | 85.6 |
| European | rs77542162 | G | A | 0.020 | 0.1831 | 0.0051 | 4.8E-212 | 1297.3 |
| European | rs72631343 | G | C | 0.130 | -0.0439 | 0.0021 | 3.7E-75 | 452.0 |
| European | rs2125345 | C | T | 0.292 | -0.0167 | 0.0015 | 4.0E-21 | 119.5 |
| European | rs12451056 | T | C | 0.144 | -0.0189 | 0.0020 | 9.8E-17 | 92.7 |
| European | rs2840354 | T | C | 0.191 | 0.0116 | 0.0018 | 2.0E-08 | 42.3 |
| European | rs77960347 | G | A | 0.013 | 0.0699 | 0.0062 | 2.2E-22 | 127.3 |
| European | rs10438978 | T | C | 0.180 | -0.0178 | 0.0018 | 2.9E-17 | 95.9 |
| European | rs12968116 | T | C | 0.126 | 0.0142 | 0.0021 | 4.9E-09 | 46.0 |
| European | rs1135908 | T | G | 0.146 | 0.0131 | 0.0020 | 2.0E-08 | 42.4 |
| European | rs4807570 | A | G | 0.210 | -0.0115 | 0.0017 | 5.8E-09 | 45.5 |
| European | rs112313064 | C | T | 0.370 | 0.0099 | 0.0016 | 3.6E-08 | 40.8 |
| European | rs1808664 | G | A | 0.394 | 0.0236 | 0.0014 | 5.3E-45 | 266.2 |
| European | rs73015007 | A | G | 0.241 | -0.0757 | 0.0016 | <4.9E-324 | 2127.3 |
| European | rs11670740 | G | A | 0.234 | -0.0266 | 0.0017 | 2.0E-43 | 256.5 |
| European | rs440677 | G | A | 0.381 | 0.0209 | 0.0015 | 5.5E-35 | 204.5 |
| European | rs62120366 | A | C | 0.350 | 0.0096 | 0.0015 | 1.4E-08 | 43.3 |
| European | rs150057262 | G | C | 0.011 | -0.1627 | 0.0073 | 7.1E-82 | 493.4 |
| European | rs17217098 | A | G | 0.067 | -0.0984 | 0.0028 | 7.0E-204 | 1246.9 |
| European | rs147791730 | A | G | 0.028 | -0.0291 | 0.0042 | 1.9E-09 | 48.5 |
| European | rs78242215 | A | G | 0.056 | -0.0207 | 0.0030 | 4.2E-09 | 46.4 |
| European | rs62119267 | C | A | 0.021 | -0.2248 | 0.0049 | <4.9E-324 | 2082.2 |

| | | | | | | | | |
|---|---|---|---|---|---|---|---|---|
| European | rs531660643 | T | G | 0.019 | -0.3927 | 0.0057 | <4.9E-324 | 4799.6 |
| European | rs140365836 | A | G | 0.006 | 0.1277 | 0.0125 | 1.2E-18 | 104.4 |
| European | rs10426226 | A | G | 0.042 | -0.0311 | 0.0035 | 3.2E-14 | 77.4 |
| European | rs113330691 | A | G | 0.034 | -0.2232 | 0.0038 | <4.9E-324 | 3405.3 |
| European | rs148601586 | G | C | 0.012 | 0.1617 | 0.0067 | 7.0E-97 | 586.0 |
| European | rs41289514 | G | A | 0.010 | 0.2509 | 0.0071 | 7.8E-204 | 1246.6 |
| European | rs72654437 | A | G | 0.030 | 0.0626 | 0.0045 | 6.6E-33 | 191.8 |
| European | rs12691088 | A | G | 0.026 | 0.1779 | 0.0047 | 3.5E-231 | 1415.6 |
| European | rs150554721 | T | C | 0.019 | -0.0384 | 0.0054 | 8.5E-10 | 50.6 |
| European | rs4803818 | T | C | 0.025 | 0.0291 | 0.0045 | 2.9E-08 | 41.3 |
| European | rs2617801 | C | G | 0.401 | 0.0215 | 0.0015 | 3.9E-36 | 211.6 |
| European | rs144794875 | G | A | 0.142 | -0.0177 | 0.0023 | 3.2E-11 | 59.1 |
| European | rs35081008 | T | C | 0.155 | -0.0309 | 0.0019 | 2.9E-43 | 255.5 |
| European | rs11673465 | C | T | 0.210 | 0.0163 | 0.0017 | 7.0E-16 | 87.5 |
| European | rs73075609 | T | C | 0.024 | 0.0479 | 0.0048 | 3.3E-18 | 101.7 |
| European | rs969075 | T | C | 0.335 | -0.0147 | 0.0015 | 2.2E-17 | 96.6 |
| European | rs2618566 | G | T | 0.339 | 0.0402 | 0.0015 | 1.1E-116 | 708.2 |
| European | rs1044573 | G | A | 0.495 | 0.0103 | 0.0014 | 3.1E-10 | 53.2 |
| European | rs224424 | G | A | 0.215 | -0.0235 | 0.0017 | 3.3E-33 | 193.6 |
| European | rs1883711 | C | G | 0.032 | 0.1308 | 0.0042 | 6.3E-161 | 981.5 |
| European | rs35570186 | A | G | 0.038 | 0.0264 | 0.0039 | 7.0E-09 | 45.0 |
| European | rs6093446 | A | G | 0.275 | 0.0242 | 0.0016 | 8.3E-41 | 240.3 |
| European | rs1800961 | T | C | 0.034 | -0.0549 | 0.0039 | 7.6E-35 | 203.7 |
| European | rs389877 | G | C | 0.191 | -0.0122 | 0.0018 | 8.0E-09 | 44.7 |
| European | rs1569750 | G | A | 0.306 | 0.0113 | 0.0015 | 2.2E-10 | 54.1 |
| European | rs6022851 | T | C | 0.446 | 0.0116 | 0.0014 | 2.3E-12 | 66.1 |
| European | rs117590445 | T | C | 0.016 | -0.0379 | 0.0059 | 2.6E-08 | 41.6 |
| European | rs2257885 | A | G | 0.247 | 0.0123 | 0.0016 | 6.6E-11 | 57.3 |
| European | rs6090040 | A | C | 0.483 | 0.0143 | 0.0015 | 4.8E-17 | 94.6 |
| European | rs73147887 | G | C | 0.212 | 0.0159 | 0.0018 | 1.9E-14 | 78.7 |
| European | rs12106385 | A | T | 0.019 | -0.0364 | 0.0055 | 1.2E-08 | 43.6 |
| European | rs112244600 | T | C | 0.051 | 0.0301 | 0.0036 | 3.1E-13 | 71.4 |
| European | rs1963676 | C | T | 0.424 | -0.0144 | 0.0014 | 4.2E-18 | 101.1 |
| European | rs5746498 | C | T | 0.238 | 0.0133 | 0.0017 | 1.4E-11 | 61.4 |
| European | rs8139142 | C | T | 0.200 | -0.0135 | 0.0020 | 7.9E-09 | 44.7 |
| European | rs5752963 | A | G | 0.037 | 0.0268 | 0.0037 | 2.7E-10 | 53.6 |

| | | | | | | | | |
|---|---|---|---|---|---|---|---|---|
| European | rs5755688 | A | G | 0.359 | 0.0137 | 0.0015 | 5.6E-16 | 88.1 |
| European | rs138352 | T | G | 0.346 | 0.0138 | 0.0015 | 7.6E-16 | 87.3 |
| European | rs9615108 | C | G | 0.387 | -0.0098 | 0.0014 | 5.6E-09 | 45.6 |
| European | rs13268 | G | A | 0.024 | -0.0380 | 0.0045 | 4.4E-13 | 70.4 |
| East Asian | rs151193009 | T | C | 0.009 | -0.4860 | 0.0333 | 2.3E-44 | 212.7 |
| East Asian | rs72911441 | G | A | 0.054 | 0.0773 | 0.0111 | 2.5E-11 | 48.5 |
| East Asian | rs6663252 | C | T | 0.120 | -0.0751 | 0.0078 | 3.5E-20 | 92.2 |
| East Asian | rs602633 | T | G | 0.066 | -0.1919 | 0.0104 | 2.4E-69 | 337.6 |
| East Asian | rs2642438 | A | G | 0.177 | -0.0509 | 0.0068 | 8.9E-13 | 55.7 |
| East Asian | rs11320208 | A | C | 0.256 | -0.0402 | 0.0057 | 1.4E-11 | 49.8 |
| East Asian | rs13306194 | A | G | 0.114 | -0.1235 | 0.0079 | 8.6E-51 | 244.8 |
| East Asian | rs62133263 | C | G | 0.076 | 0.0696 | 0.0095 | 2.0E-12 | 53.9 |
| East Asian | rs2539981 | T | C | 0.328 | -0.0373 | 0.0055 | 7.6E-11 | 46.2 |
| East Asian | rs6741916 | T | A | 0.271 | -0.0414 | 0.0063 | 3.8E-10 | 42.7 |
| East Asian | rs3752442 | G | A | 0.453 | -0.0330 | 0.0054 | 4.7E-09 | 37.4 |
| East Asian | rs6453131 | T | G | 0.498 | -0.0815 | 0.0050 | 4.3E-55 | 266.3 |
| East Asian | rs6874202 | T | C | 0.253 | -0.0557 | 0.0058 | 2.3E-20 | 93.2 |
| East Asian | rs9376090 | C | T | 0.280 | -0.0398 | 0.0057 | 2.6E-11 | 48.4 |
| East Asian | rs73596816 | A | G | 0.032 | 0.1160 | 0.0141 | 2.8E-15 | 68.0 |
| East Asian | rs188951644 | A | T | 0.003 | 0.3689 | 0.0506 | 2.8E-12 | 53.2 |
| East Asian | rs7789194 | T | C | 0.421 | 0.0430 | 0.0055 | 9.3E-14 | 60.5 |
| East Asian | rs2054345 | C | T | 0.012 | -0.1352 | 0.0236 | 4.2E-08 | 32.7 |
| East Asian | rs112784971 | T | C | 0.244 | -0.0363 | 0.0059 | 2.8E-09 | 38.5 |
| East Asian | rs2954027 | T | A | 0.451 | 0.0432 | 0.0051 | 3.5E-16 | 72.4 |
| East Asian | rs1883025 | T | C | 0.234 | -0.0397 | 0.0059 | 9.8E-11 | 45.6 |
| East Asian | rs59857465 | A | G | 0.076 | 0.0563 | 0.0098 | 3.9E-08 | 32.9 |
| East Asian | rs2519093 | T | C | 0.216 | 0.0860 | 0.0060 | 1.1E-42 | 204.3 |
| East Asian | rs7898735 | C | T | 0.238 | -0.0578 | 0.0061 | 1.8E-19 | 88.8 |
| East Asian | rs2419607 | G | T | 0.403 | -0.0366 | 0.0052 | 1.0E-11 | 50.4 |
| East Asian | rs11601507 | A | C | 0.084 | 0.0597 | 0.0093 | 6.8E-10 | 41.5 |
| East Asian | rs174559 | A | G | 0.410 | -0.0358 | 0.0056 | 8.9E-10 | 40.9 |
| East Asian | rs59379014 | T | C | 0.094 | 0.0538 | 0.0088 | 5.0E-09 | 37.3 |
| East Asian | rs12229026 | C | T | 0.282 | 0.0347 | 0.0057 | 6.0E-09 | 36.9 |
| East Asian | rs4646776 | C | G | 0.195 | 0.0390 | 0.0064 | 6.4E-09 | 36.7 |
| East Asian | rs11571836 | G | A | 0.376 | -0.0429 | 0.0052 | 4.4E-15 | 67.0 |
| East Asian | rs7140110 | C | T | 0.217 | 0.0430 | 0.0067 | 9.9E-10 | 40.7 |

| | | | | | | | | |
|---|---|---|---|---|---|---|---|---|
| East Asian | rs6493583 | G | C | 0.174 | -0.0410 | 0.0069 | 1.6E-08 | 34.8 |
| East Asian | rs77303550 | T | C | 0.244 | -0.0895 | 0.0064 | 5.3E-41 | 195.9 |
| East Asian | rs7212349 | T | C | 0.274 | -0.0340 | 0.0058 | 1.4E-08 | 35.0 |
| East Asian | rs62074055 | C | G | 0.332 | 0.0382 | 0.0055 | 2.6E-11 | 48.4 |
| East Asian | rs12162136 | G | A | 0.400 | 0.0341 | 0.0051 | 1.4E-10 | 44.9 |
| East Asian | rs2738464 | G | C | 0.283 | -0.1080 | 0.0059 | 8.3E-68 | 329.9 |
| East Asian | rs737337 | C | T | 0.265 | -0.0619 | 0.0059 | 5.0E-24 | 111.3 |
| East Asian | rs11668738 | G | T | 0.421 | -0.0445 | 0.0060 | 1.7E-12 | 54.2 |
| East Asian | rs7254892 | A | G | 0.053 | -0.5976 | 0.0126 | <4.9E-324 | 2241.5 |
| East Asian | rs187976859 | C | A | 0.005 | -0.3052 | 0.0400 | 2.6E-13 | 58.3 |
| East Asian | rs5758600 | T | G | 0.433 | 0.0439 | 0.0057 | 1.2E-13 | 59.9 |

LDL, low-density lipoprotein.

Supplemental Table 5. Variance in plasma LDL-cholesterol explained by SNPs for each therapy and plasma LDL-cholesterol in people of European and East Asian ancestry.

| Ancestry | Exposure | Gene | Number of SNPs | Variance explained |
|---|---|---|---|---|
| European | Statins | *HMGCR* | 21 | 0.5% |
| European | PCSK9 inhibitors | *PCSK9* | 33 | 1.5% |
| European | Ezetimibe | *NPC1L1* | 6 | 0.1% |
| European | Targeting LDL receptors | *LDLR* | 42 | 2.4% |
| European | Mipomersen | *APOB* | 25 | 1.7% |
| European | Targeting ABCG5/8 | *ABCG5/8* | 25 | 0.7% |
| European | Plasma LDL-cholesterol | Across the genome | 324 | 8.7% |
| East Asian | Statins | *HMGCR* | 5 | 0.6% |
| East Asian | PCSK9 inhibitors | *PCSK9* | 7 | 0.9% |
| East Asian | Targeting LDL receptors | *LDLR* | 9 | 1.2% |
| East Asian | Mipomersen | *APOB* | 5 | 0.6% |
| East Asian | Plasma LDL-cholesterol | Across the genome | 43 | 8.6% |

ABCG5/8, adenosine triphosphate (ATP)-binding cassette transporters G5/8; LDL, low-density lipoprotein; PCSK9, proprotein convertase subtilisin/kexin type 9.

Supplemental Table 6. SNPs associated with common confounders at genome-wide significant level (*p* value <5.0E-8) in PhenoScanner.

| SNP | Trait | Author | Study/PMID | Ancestry | *P* value |
|---|---|---|---|---|---|
| rs11499828 | Alcohol intake frequency | Neale B | UK Biobank | European | 3.8E-17 |
| rs174547 | Average weekly champagne plus white wine intake | Neale B | UK Biobank | European | 2.6E-08 |
| rs3184504 | Past tobacco smoking | Neale B | UK Biobank | European | 5.5E-11 |
| rs3184504 | Smoking status: previous | Neale B | UK Biobank | European | 6.2E-10 |
| rs4646776 | Alcohol consumption | Yang X | 23364009 | East Asian | 5.2E-35 |

Supplemental Table 7. Colocalization estimates for each posterior probability using prior probabilities 1.0E-4 for a variant associated with plasma LDL-cholesterol, 1.0E-4 for a variant associated with gallstone disease, and 1.0E-5 for a variant associated with both traits in or near (+-100kb) the target gene of each therapy.

| Therapy | Gene | Biobank | SNPs | $H_0$ | $H_1$ | $H_2$ | $H_3$ | $H_4$ | Conditional $H_4$ |
|---|---|---|---|---|---|---|---|---|---|
| Statins | *HMGCR* | FinnGen | 877 | <0.001 | 0.001 | <0.001 | 0.012 | 0.987 | 0.988 |
| PCSK9 inhibitors | *PCSK9* | UK Biobank | 1298 | <0.001 | 0.912 | <0.001 | 0.057 | 0.031 | 0.352 |
| Mipomersen | *APOB* | UK Biobank | 1085 | <0.001 | 0.504 | <0.001 | 0.447 | 0.049 | 0.099 |
| Mipomersen | *APOB* | FinnGen | 930 | <0.001 | 0.944 | <0.001 | 0.042 | 0.014 | 0.247 |
| Targeting ABCG5/8 | *ABCG5/8* | UK Biobank | 1862 | <0.001 | <0.001 | <0.001 | >0.999 | <0.001 | <0.001 |
| Targeting ABCG5/8 | *ABCG5/8* | FinnGen | 1389 | <0.001 | <0.001 | <0.001 | >0.999 | <0.001 | <0.001 |

ABCG5/8, adenosine triphosphate (ATP)-binding cassette transporters G5/8; LDL, low-density lipoprotein; PCSK9, proprotein convertase subtilisin/kexin type 9. $H_0$, no association with either trait; $H_1$, association with plasma LDL-cholesterol only; $H_2$, association with gallstone disease only; $H_3$, associations of two independent variants and one for each trait; $H_4$, associations of one shared variant with both traits; conditional $H_4$, associations of one shared variant with both traits conditional on the presence of a variant associated with gallstone disease ($H_4/(H_2+H_3+H_4)$).

Supplemental Table 8. Cluster-specific SNPs with inclusion probability >0.80 in the UK Biobank and FinnGen.

| Cluster | Number of overlapping SNPs | Number of SNPs in UK Biobank cluster only | Number of SNPs in FinnGen cluster only | Overlapping SNPs | SNPs in UK Biobank cluster only | SNPs in FinnGen cluster only |
|---|---|---|---|---|---|---|
| Cluster1 | 3 | 2 | 3 | rs1497406: rs1896995: rs9297994 | rs2257885: rs2617801 | rs13108218: rs17580: rs352942 |
| Cluster2 | 4 | 12 | 6 | rs11167778: rs2519093: rs3010275: rs67890964 | rs11621792: rs11670740: rs12533280: rs13108218: rs13379043: rs17580: rs1801689: rs2642438: rs440677: rs7300192: rs7904973: rs9837622 | rs10272002: rs114165349: rs1423527: rs2125345: rs2617801: rs4671050 |
| Cluster3 | 11 | 21 | 1 | rs10455872: rs113330691: rs12691088: rs35271870: rs35358959: rs41289514: rs531660643: rs62119267: rs62122481: rs693668: rs9987289 | rs11206517: rs11591147: rs11601507: rs117733303: rs11787335: rs12208357: rs12603290: rs12657266: rs13076933: rs17050272: rs17217098: rs1808664: rs247617: rs34042070: rs34707604: rs553427: rs6689: rs71311871: rs73015007: rs77542162: rs9399137 | rs72631343 |

| | | | | | | |
|---|---|---|---|---|---|---|
| Cluster4 | 4 | 1 | 17 | rs10903129: rs174547: rs2618566: rs28601761 | rs55714927 | rs1057868: rs11673465: rs1169288: rs12445804: rs12917376: rs138204164: rs1556857: rs2391159: rs267733: rs2706381: rs35081008: rs4390169: rs5752963: rs603424: rs6792725: rs799157: rs9496567 |
| Cluster5 | 3 | 1 | 1 | rs12968116: rs1800961: rs6741740 | rs185263492 | rs2374569 |

Supplemental Table 9. KEGG and Reactome pathways related to the mapped genes for cluster-specific variants predicting plasma LDL-cholesterol (inclusion probability >0.80).

| Biobank | Cluster | Category | Pathway | Number of genes of the pathway | Number of input genes overlapping with genes of the pathway | Input genes overlapping with genes of the pathway | *P* value | Adjusted *p* value |
|---|---|---|---|---|---|---|---|---|
| FinnGen | Cluster2 | KEGG | Glycosphingolipid biosynthesis globo series | 14 | 3 | *FUT2:FUT1:GBGT1* | 3.0E-05 | 5.5E-03 |
| FinnGen | Cluster3 | KEGG | ABC transporters | 42 | 4 | *ABCA9:ABCA6:ABCA10:ABCA5* | 3.1E-06 | 5.7E-04 |
| FinnGen | Cluster3 | Reactome | Transport of small molecules | 721 | 14 | *BSND:PCSK9:PSMA5:ABCA9:ABCA6:ABCA10:ABCA5:APOE:APOC1:APOC4:APOC2:SLC22A2:SLC22A3:LPA* | 8.0E-10 | 1.2E-06 |
| FinnGen | Cluster3 | Reactome | Plasma lipoprotein assembly remodeling and clearance | 71 | 6 | *PCSK9:APOE:APOC1:APOC4:APOC2:LPA* | 1.7E-08 | 1.3E-05 |
| FinnGen | Cluster3 | Reactome | ABC transporters in lipid homeostasis | 18 | 4 | *ABCA9:ABCA6:ABCA10:ABCA5* | 8.7E-08 | 4.1E-05 |
| FinnGen | Cluster3 | Reactome | Plasma lipoprotein assembly | 19 | 4 | *APOE:APOC1:APOC4:APOC2* | 1.1E-07 | 4.1E-05 |
| FinnGen | Cluster3 | Reactome | Plasma lipoprotein clearance | 33 | 4 | *PCSK9:APOE:APOC1:APOC4* | 1.1E-06 | 3.4E-04 |

| | | | | | | | | |
|---|---|---|---|---|---|---|---|---|
| FinnGen | Cluster3 | Reactome | ABC family proteins mediated transport | 99 | 5 | *PSMA5:ABCA9:ABCA6:ABCA10:ABCA5* | 3.9E-06 | 9.7E-04 |
| FinnGen | Cluster3 | Reactome | Abacavir transmembrane transport | 5 | 2 | *SLC22A2:SLC22A3* | 5.6E-05 | 1.0E-02 |
| FinnGen | Cluster3 | Reactome | VLDL assembly | 5 | 2 | *APOC1:APOC4* | 5.6E-05 | 1.0E-02 |
| FinnGen | Cluster3 | Reactome | Plasma lipoprotein remodeling | 32 | 3 | *APOE:APOC2:LPA* | 6.1E-05 | 1.0E-02 |
| FinnGen | Cluster3 | Reactome | VLDL clearance | 6 | 2 | *APOC1:APOC4* | 8.4E-05 | 1.3E-02 |
| FinnGen | Cluster3 | Reactome | Abacavir transport and metabolism | 10 | 2 | *SLC22A2:SLC22A3* | 2.5E-04 | 2.5E-02 |
| FinnGen | Cluster3 | Reactome | Organic cation transport | 10 | 2 | *SLC22A2:SLC22A3* | 2.5E-04 | 2.5E-02 |
| FinnGen | Cluster3 | Reactome | Chylomicron assembly | 10 | 2 | *APOE:APOC2* | 2.5E-04 | 2.5E-02 |
| FinnGen | Cluster3 | Reactome | Chylomicron remodeling | 10 | 2 | *APOE:APOC2* | 2.5E-04 | 2.5E-02 |
| FinnGen | Cluster3 | Reactome | HDL remodeling | 10 | 2 | *APOE:APOC2* | 2.5E-04 | 2.5E-02 |
| UK Biobank | Cluster3 | Reactome | Plasma lipoprotein assembly remodeling and clearance | 71 | 7 | *PCSK9:CETP:APOE:APOC1:APOC4:APOC2:LPA* | 2.3E-06 | 3.5E-03 |
| UK Biobank | Cluster3 | Reactome | Abacavir transmembrane transport | 5 | 3 | *SLC22A1:SLC22A2:SLC22A3* | 5.9E-06 | 4.4E-03 |

| | | | | | | | | |
|---|---|---|---|---|---|---|---|---|
| UK Biobank | Cluster3 | Reactome | Transport of small molecules | 721 | 19 | *BSND:PCSK9:PSMA5:SLC12A3:CETP:ABCA9:ABCA6:ABCA10:ABCA5:SLC44A2:ATP13A1:APOE:APOC1:APOC4:APOC2:SLC22A1:SLC22A2:SLC22A3:LPA* | 1.2E-05 | 5.2E-03 |
| UK Biobank | Cluster3 | Reactome | ABC transporters in lipid homeostasis | 18 | 4 | *ABCA9:ABCA6:ABCA10:ABCA5* | 1.4E-05 | 5.2E-03 |
| UK Biobank | Cluster3 | Reactome | Plasma lipoprotein assembly | 19 | 4 | *APOE:APOC1:APOC4:APOC2* | 1.7E-05 | 5.2E-03 |
| UK Biobank | Cluster3 | Reactome | Abacavir transport and metabolism | 10 | 3 | *SLC22A1:SLC22A2:SLC22A3* | 6.8E-05 | 1.3E-02 |
| UK Biobank | Cluster3 | Reactome | Organic cation transport | 10 | 3 | *SLC22A1:SLC22A2:SLC22A3* | 6.8E-05 | 1.3E-02 |
| UK Biobank | Cluster3 | Reactome | HDL remodeling | 10 | 3 | *CETP:APOE:APOC2* | 6.8E-05 | 1.3E-02 |
| UK Biobank | Cluster3 | Reactome | Plasma lipoprotein remodeling | 32 | 4 | *CETP:APOE:APOC2:LPA* | 1.5E-04 | 2.5E-02 |
| UK Biobank | Cluster3 | Reactome | Plasma lipoprotein clearance | 33 | 4 | *PCSK9:APOE:APOC1:APOC4* | 1.7E-04 | 2.5E-02 |
| UK Biobank | Cluster3 | Reactome | Organic cation anion zwitterion transport | 15 | 3 | *SLC22A1:SLC22A2:SLC22A3* | 2.5E-04 | 3.4E-02 |
| Biobank Japan | Cluster2 | Reactome | Plasma lipoprotein assembly remodeling and clearance | 71 | 10 | *PCSK9:LDLR:C19orf80:APOE:APOC1:APOC4:APOC2:APOB:LPA:ABCA1* | 1.2E-10 | 1.8E-07 |

| | | | | | | | | |
|---|---|---|---|---|---|---|---|---|
| Biobank Japan | Cluster2 | Reactome | Plasma lipoprotein assembly | 19 | 6 | *APOE:APOC1:APOC4:APOC2:APOB:ABCA1* | 3.9E-09 | 2.9E-06 |
| Biobank Japan | Cluster2 | Reactome | Plasma lipoprotein clearance | 33 | 6 | *PCSK9:LDLR:APOE:APOC1:APOC4:APOB* | 1.5E-07 | 7.3E-05 |
| Biobank Japan | Cluster2 | Reactome | Transport of small molecules | 721 | 19 | *BSND:PCSK9:MICU1:MCU:BEST1:FTH1:SLC44A2:LDLR:C19orf80:APOE:APOC1:APOC4:APOC2:APOB:ADD1:SLC22A3:LPA:ABCA1:SLC44A1* | 1.8E-06 | 6.8E-04 |
| Biobank Japan | Cluster2 | Reactome | Plasma lipoprotein remodeling | 32 | 5 | *C19orf80:APOE:APOC2:APOB:LPA* | 3.7E-06 | 8.6E-04 |
| Biobank Japan | Cluster2 | Reactome | VLDL assembly | 5 | 3 | *APOC1:APOC4:APOB* | 4.0E-06 | 8.6E-04 |
| Biobank Japan | Cluster2 | Reactome | Chylomicron clearance | 5 | 3 | *LDLR:APOE:APOB* | 4.0E-06 | 8.6E-04 |
| Biobank Japan | Cluster2 | Reactome | VLDL clearance | 6 | 3 | *APOC1:APOC4:APOB* | 8.0E-06 | 1.5E-03 |
| Biobank Japan | Cluster2 | Reactome | Chylomicron assembly | 10 | 3 | *APOE:APOC2:APOB* | 4.7E-05 | 7.1E-03 |
| Biobank Japan | Cluster2 | Reactome | Chylomicron remodeling | 10 | 3 | *APOE:APOC2:APOB* | 4.7E-05 | 7.1E-03 |
| Biobank Japan | Cluster2 | Reactome | Visual phototransduction | 99 | 6 | *GRK1:LDLR:APOE:APOC2:APOB:GRK4* | 9.9E-05 | 1.4E-02 |
| Biobank Japan | Cluster2 | Reactome | Regulation of IGF transport and uptake by IGFBPs | 123 | 6 | *PCSK9:TMEM132A:GAS6:APOE:APOB:PLG* | 3.2E-04 | 3.9E-02 |

| | | | | | | | | |
|---|---|---|---|---|---|---|---|---|
| Biobank Japan | Cluster2 | Reactome | Scavenging by class A receptors | 19 | 3 | *FTH1:APOE:APOB* | 3.6E-04 | 3.9E-02 |
| Biobank Japan | Cluster2 | Reactome | LDL clearance | 19 | 3 | *PCSK9:LDLR:APOB* | 3.6E-04 | 3.9E-02 |
| Biobank Japan | Cluster2 | Reactome | Metabolism of fat soluble vitamins | 47 | 4 | *LDLR:APOE:APOC2:APOB* | 4.2E-04 | 4.1E-02 |

ABC, adenosine triphosphate (ATP)-binding cassette transporters; HDL, high-density lipoprotein; IGF, insulin-like growth factor; IGFBPs, insulin-like growth factor-binding proteins; LDL, low-density lipoprotein; VLDL, very-low-density lipoprotein.
